# Multi-site disease analytics with applications to estimating COVID-19 undetected cases in Canada

**DOI:** 10.1101/2022.07.11.22277508

**Authors:** Matthew R. P. Parker, Jiguo Cao, Laura L. E. Cowen, Lloyd T. Elliott, Junling Ma

**Affiliations:** Department of Statistics and Actuarial Science, Simon Fraser University, Canada; Department of Mathematics and Statistics, University of Victoria, Canada

## Abstract

Even with daily case counts, the true scope of the COVID-19 pandemic in Canada is unknown due to undetected cases. We estimate the pandemic scope through a new multi-site model using publicly available disease count data including detected cases, recoveries among detected cases, and total deaths. These counts are used to estimate the case detection probability, the infection fatality rate through time, as well as the probability of recovery, and several important population parameters including the rate of spread, and importation of external cases. We also estimate the total number of active COVID-19 cases per region of Canada for each reporting interval. We applied this multi-site model Canada-wide to all provinces and territories, providing an estimate of the total COVID-19 burden for the 90 weeks from 23 Apr 2020 to 6 Jan 2022. We also applied this model to the five Health Authority regions of British Columbia, Canada, describing the pandemic in B.C. over the 31 weeks from 2 Apr 2020 to 30 Oct 2020.

## 1 Introduction

The novel coronavirus pandemic began in December 2019, and spread rapidly to every corner of the globe. The impact of the pandemic on lives, societal and social norms, and on the world economy cannot be overstated (Béland et al., 2021; Cypress, 2022; Tanaka, 2022). As of late June 2022, the global total number of deaths attributed to the pandemic is over 6.3 million, from a recorded total of 540 million reported cases. And Canada has recorded a total of over 3.9 million confirmed cases, and over 41 thousand confirmed deaths (Dong et al., 2020). The number of confirmed cases of coronavirus are an under-reporting of the true total (see for example: Feikin et al., 2020; Buitrago-Garcia et al., 2020; Hasan et al., 2021; Chisale et al., 2022). The reasons for under-reporting are varied and include the presence of asymptomatic, pauci-symptomatic, and pre-symptomatic cases, lack of testing for low severity cases, and periods with low testing volumes. Some of these causes can be controlled through testing protocols and wider availability/accessibility of testing. However, due to the impracticality of full census testing of large populations in Canada, under-reporting cannot be entirely mitigated. Undetected cases cause community infections, and reduces the effectiveness of control measures such as contact tracing, quarantine and isolation. They also cause underestimates of the social and economical impacts of the pandemic.

To properly understand the scope and impact of the pandemic, we must estimate the true total number of infections. Research conducted after the pandemic began has improved methodologies to produce such estimates. These methods include meta-analyses of asymptomatic prevalence (He et al., 2021; Alene et al., 2021), extensions to susceptible-infectious-recovered type modelling (Li et al., 2021; Subramanian et al., 2021; Huo et al., 2021), and seroprevalence studies (Bendavid et al., 2021; Saeed et al., 2021; Halili et al., 2022). Alternative models include integer-autoregressive models (Fernández-Fontelo et al., 2020) and case fatality rate (CFR) models (Dougherty et al., 2021), and estimating under-reporting using discrete count models (Parker et al., 2021).

Much research has focused on understanding the course of the pandemic within Canada. Examples include seroprevalence of Montréal school age children (Zinszer et al., 2021), CFR based models to estimate reporting rates across Canada (Dougherty et al., 2021), intervention strategy analysis in Ontario (Tuite et al., 2020), mental health and well-being of Canadians during the pandemic (Dozois, 2021; Appleby et al., 2022), models to forecast transmission and incidence (Chimmula and Zhang, 2020; Mullah and Yan, 2022), and policy response analysis with comparisons between Canada, France, and Belgium (Desson et al., 2020). Among this backdrop of vital research, we provide up-to-date disease analytics for Canada as a whole, with results specific to each province and territory.

We propose a novel multi-site modelling technique to better estimate disease dynamics such as domestic spread rate, recovery and death probabilities, and to estimate effectiveness of testing protocols and levels of under-reporting of cases. By considering data from across the entire time span of the pandemic, estimates can be made throughout its course. Multi-site modelling allows the total burden of the pandemic to be estimated across large regions by considering them as an amalgamation of smaller regions. This method of modelling increases precision compared to single site modelling, by pooling information across sites, which is an example of transfer learning (Weiss et al., 2016). To analyze the burden of COVID-19 as it has progressed through time in Canada, we model the entire pandemic period to date (from 23 Apr 2020 to 6 Jan 2022).

Our multi-site hidden Markov model has several new mathematical contributions in comparison with the single-site model (Parker et al., 2021): We provide a multi-site framework for disease analytics, we add a saturation effect to model limitations on disease spread, and we model the disease spread rate separately for the detected and the undetected disease cases. The latent states in our model are identifiable due to parametric assumptions. This is similar to the modelling of population abundance in open population N-mixtures models (Dail and Madsen, 2011).

We investigate the effect of vaccination coverage on virus spread rates, recovery probability, and death probability through inclusion of a vaccine coverage covariate. We also include indicators for time periods demarcated by the first confirmed cases in Canada of the variants of concern Delta (Mahumud et al., 2022) and Omicron (Araf et al., 2022), which have been shown to have very different vaccine efficacies (Kahn et al., 2022). These indicators allow us to model changes in disease dynamics and interaction between vaccine efficacy and the dominant variant of concern.

The main contributions of this paper are (1) a novel multi-site disease analytics model, (2) estimates of the total burden of COVID-19 across Canada for 90 weeks of the pandemic, (3) estimates of reporting rates for each province and territory of Canada, (4) estimates of domestic spread rates among both detected and undetected cases, (5) estimates of infection fatality rate (IFR) for COVID-19 in Canada, (6) estimates of average recovery period for active cases, and (7) comparisons between competing models.

## 2 Methods

### 2.1 Multi-site Model

In our new multiple-site disease analytics model, each site is treated as statistically independent, so that the likelihood function is a product of single-site likelihoods. Figure 1 outlines the single-site model including the disease dynamics (top panel), detection mechanisms (bottom left panel), and the difference between the model definition of the data and the actual reporting times (bottom right panel).

**Figure 1:**
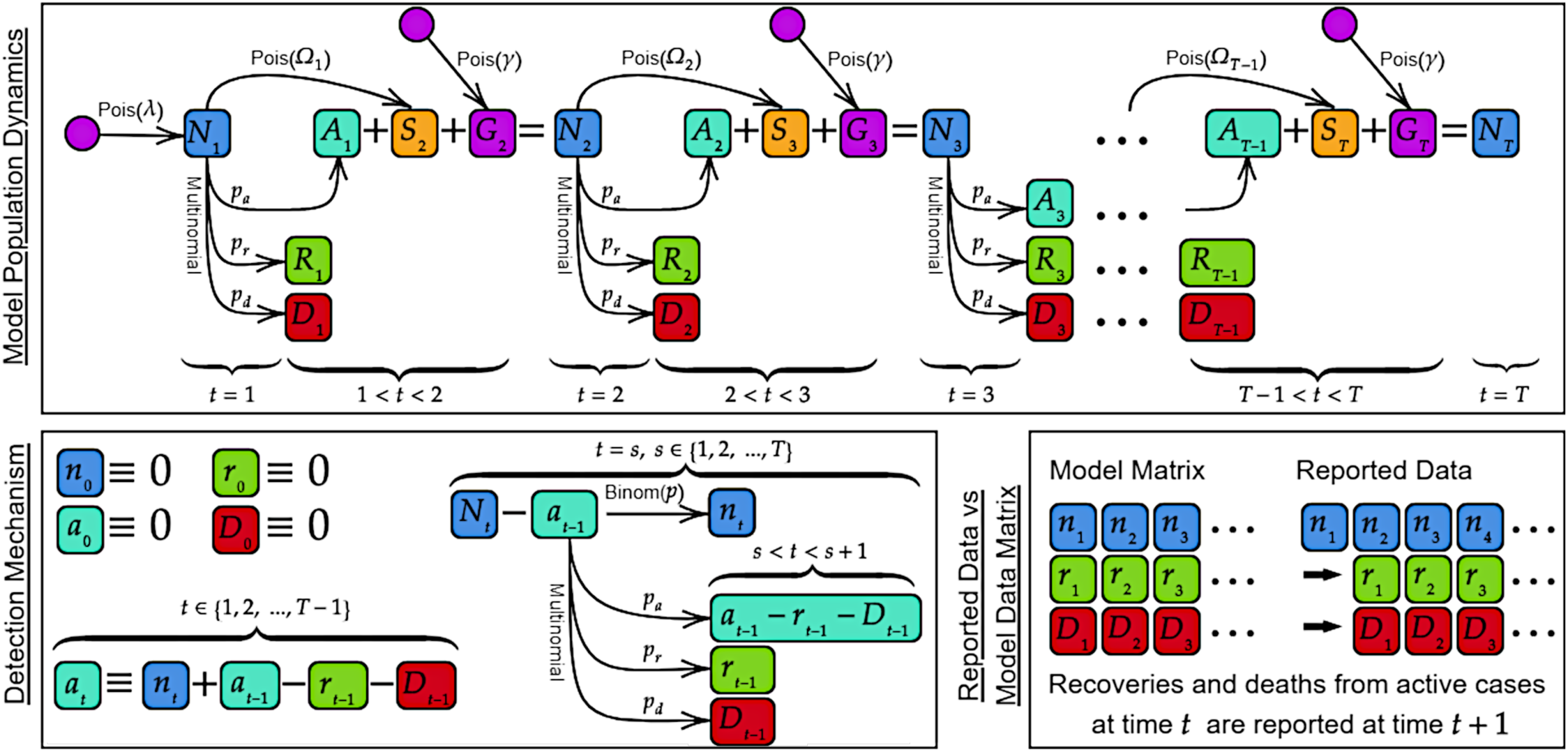
Diagram of single-site model. Top: The disease dynamics and state process. Bottom left: The detection mechanisms. Bottom right: Data reporting times.

Let *n*_*it*_ denote the new detected case counts at sampling occasion *t* = 1, …, *T* and study site *i* = 1, …, *M*. The new detected recoveries between *t* and *t* + 1 among all detected active cases are *r*_*it*_. The new detected deaths between *t* and *t* + 1 are *d*_*it*_ (when deaths are fully detected, we let *d*_*it*_ = *D*_*it*_, and *D*_*it*_ cease to be considered latent states). The total number of active cases at time *t* are *N*_*it*_, and *a*_*it*_ cases among them are detected. The active cases at time *t* that will recover, die, or remain active between *t* and *t* + 1 are respectively *R*_*it*_, *D*_*it*_, *A*_*it*_. In the following we split the model into ten components for ease of understanding:

1. Initial Abundance: *N*_*i*1_ ∼ Poisson(λ)
2. State Process: {*A*_*it*_, *D*_*it*_, *R*_*it*_} ∼ Multinomial(*N*_*it*_; *p*_*a*_, *p*_*d*_, *p*_*r*_)
3. Detected Active Cases: *a*_*it*_ = *n*_*it*_ + *a*_*it*−1_ − *r*_*it*−1_ − *d*_*it*−1_, for *t* > 0
4. Domestic Spread: *S*_*it*_ ∼ Poisson(Ω_*it*−1_), for *t* > 1
5. Ω_*it*−1_ : *ω*_1_(*N*_*it*−1_ − *a*_*it*−1_) · *δ*_*i*_ + *ω*_2_*a*_*it*−1_ (mean domestic spread)
6. *δ*_*i*_ : (*H*_*i*_ − *N*_*it*_) *H*_*i*_ (fraction of susceptible population. Here *H*_*i*_ is the total population size)
7. Imported Cases: *G*_*it*_ ∼ Poisson(γ), for *t* > 1
8. Abundance Updates: *N*_*it*_ = *A*_*it*−1_ + *S*_*it*_ + *G*_*it*_, for *t* > 1
9. Observation Process I: *n*_*it*_ ∼ Binomial(*N*_*it*_ − *a*_*it*−1_, *p*)
10. Observation Process II: 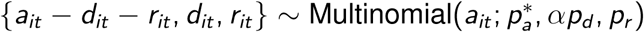

Component (1) describes the initial abundance where *λ* is defined as the expected initial population size. The latent state process (Component 2) partitions each active case in *N*_*it*_ at time *t* into one of the three categories *A*_*it*_, *D*_*it*_, and *R*_*it*_, with probabilities *p*_*a*_, *p*_*d*_, and *p*_*r*_, according to whether each case will remain active, die, or recover by time *t* + 1. Here *p*_*a*_ = 1 − *p*_*r*_ − *p*_*d*_. We assume that individuals are indistinguishable, which is a requirement for these aggregate data models, and leads to *p*_*r*_ and *p*_*d*_ being constant across individuals, and is a justification for the choice of using the multinomial distribution. Component (3) provides the calculation for *a*_*it*_ the number of detected cases still active at time *t*. The value of *a*_*it*_ is entirely specified by the observed data. We calculate *a*_*it*_ recursively, with *a*_*i*0_, *r*_*i*0_, *d*_*i*0_, *n*_*i*0_ all zero by definition, except when detected active cases are known from the time period prior to *t* = 1, in which case *a*_*i*0_ would not be zero. In Component (4), *S*_*it*_ models the domestic spread rate, which is the spread of the disease due to contact with infectious individuals within the population during each time interval. We make the simplifying assumption that the transmission rate remains constant unless mediated by additional covariates, so that the Poisson distribution is a reasonable choice. *S*_*it*_ allows for exponential growth of cases, but does not allow a spontaneous outbreak within a population having zero active cases. Ω_*it*_ is the average new infections per time interval, which is calculated using the two parameters *ω*_1_ and *ω*_2_ (Component 5). The basic reproductive number *R*_0_ is the product of Ω and the mean infectious period (1*/p*_*r*_). Parameters *ω*_1_ and *ω*_2_ are the average new infections per undetected (*N*_*it*−1_ − *a*_*it*−1_) and detected (*a*_*it*−1_) active case, respectively. Component (6) describes the value *δ*_*i*_ that modulates the growth of cases as the population becomes saturated with infection. Here *H*_*i*_ is the total population size of site *i*, which is the maximum number of infected individuals that are possible in that region. *δ*_*i*_ decreases to zero linearly as *N*_*it*_ approaches *H*_*i*_, implying that the spread rate goes to zero as the population becomes fully saturated. Imported cases, *G*_*it*_, are new cases entering the population (Component 7), for example from travel. The parameter γ is the average new number of imported cases per time interval. Imported cases allow for disease to occur even when *N*_*i*1_ = 0, allowing for spontaneous outbreaks in regions with no previous active cases. *G*_*it*_ allows for linear growth of cases over time. The number of cases *N*_*it*_ changes with time (Component 8), so we calculate the abundance updates for *t* > 1 using *A*_*it*_ from the state process as well as the growth terms *S*_*it*_ and *G*_*it*_, giving *N*_*it*_ = *A*_*it*−1_ + *S*_*it*_ + *G*_*it*_. Component (9) describes the reporting of case counts where *p* is the probability of detecting a case, and so 1 − *p* gives the under-reporting rate. We would like to have *n*_*it*_ ∼ Binomial(*N*_*it*_, *p*), which is the traditional N-mixtures parameterization of detection probability (Royle, 2004). However, since *N*_*it*_ comprises all active cases, including those which have already been detected, this would allow double counting (because detected cases *n*_*it*_ are tracked until recovery or death). Instead, we subtract the already detected active cases prior to the binomial thinning: *N*_*it*_ − *a*_*it*−1_. Individuals are indistinguishable for these aggregate count models, and so we choose the binomial distribution to model detection. Component (10) models the reporting of recoveries and deaths by partitioning *a*_*it*_ into cases that remain active, cases that die, and cases that recover. We use *α* = 1 when deaths are under-reported. In the situation where deaths are considered to be fully reported (when detected deaths are *D*_*it*_ rather than *d*_*it*_), we set *α* = 1*/p*, which increases the proportion of deaths among detected cases compared to proportion of deaths among all cases. Note that 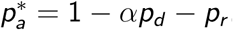.

We have built on the Parker et al. (2021) model to allow for multiple sites, which in our B.C. case study corresponds to the five Health Authority Regions of B.C., and which in our Canada-wide case study corresponds to the provinces and territories of Canada. Deaths and recoveries are occasionally recorded directly during the first observation period for an individual (such as when a recovery or death occurs in the same week as the initial positive test result). This leads to a small simplification of the detection process: *n*_*it*_ ∼ Binomial(*p, N*_*it*_ − *a*_*it*−1_) rather than *n*_*it*_ ∼ Binomial(*p, N*_*it*_ − *a*_*it*_ + *d*_*it*_ + *r*_*it*_). With this improvement, we not only continue to avoid double counting cases, but also allow deaths and recoveries to occur in the same reporting period as the first observation for an individual. We have split the spread rate *ω* into two new parameters *ω*_1_ and *ω*_2_. This allows us to model different domestic spread rates due to detected and undetected cases. To account for local cluster saturation and population saturation effects, we incorporated a penalty term to the model which allows *ω*_1_ to diminish as the number of active cases increases. By using the known population size of a region as an upper bound on total possible infections, we linearly reduce the domestic spread to zero when the population is saturated, replacing *ω*_1_(*N*_*it*_ − *a*_*it*_) with *ω*_1_(*N*_*it*_ − *a*_*it*_)(*H*_*i*_ − *N*_*it*_) *H*_*i*_. Here *H*_*i*_ is the total population of region *i*. An exponential decay could be considered rather than linear decay if case cluster saturation is expected to be a dominant effect, which can be explored in future work. The penalty term has the additional benefit of penalizing “infinite abundances”, which can be an issue for N-mixture type models (Dennis et al., 2015; Barker et al., 2018). The full joint distribution for our multi-site model is thus 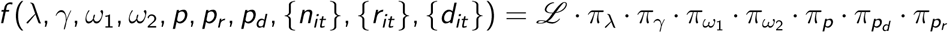 with likelihood function *L* given by the following equation:

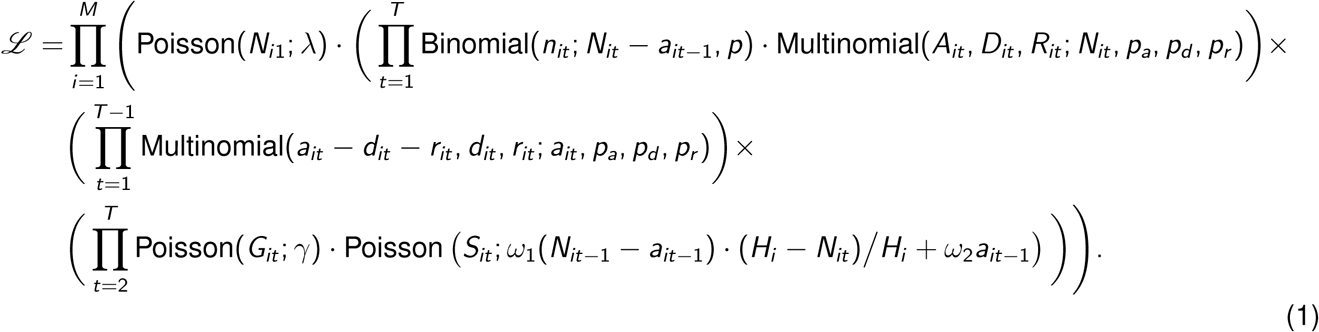

Here π_*x*_ is the prior distribution chosen for parameter *x*. Note that while the sites are independent conditioned on their parameters, the likelihood shares parameters across sites. Parameters can be separated across sites or pooled by grouping sites using categorical site covariates. A single-site model can be obtained by dropping the site subscript *i* from Equation 1 (Appendix S.1). We also conducted a simulation study to verify parameter identifiability for our multi-site model (Appendix S.2).

### 2.2 Model Fitting

We used Bayesian Markov-chain Monte-Carlo (MCMC) methods for parameter estimation and implemented model fitting using the statistical computing software R (R Core Team, 2022), together with the R package *Nimble* for probabilistic programming (de Valpine et al., 2017, 2021). We mainly used uninformative uniform prior distributions with reasonable upper and lower bounds. For example, *ω*_1_ was given a prior of Uniform(0, 5), as *ω*_1_ ≥ 0, and an *ω*_1_ of 5 is far larger than would be expected. Our investigations indicate that with the exception of extreme priors, parameter estimates were not sensitive to the prior. We found that a burn-in of 100,000 was usually enough for the MCMC chains to converge. However, for some combinations of data and covariates a burn-in of 1,000,000 was necessary. We used 200,000 iterations, which was more than sufficient to produce posterior estimates.

## 3 Applications

### 3.1 Case Study: Canada

#### 3.1.1 Data Sources

We used three publicly available sources of data for our Canada case study. The majority of the data came from the Government of Canada’s COVID-19 daily epidemiology update (PHAC: Government of Canada, 2022a), which contains testing volumes, case counts, recoveries, and deaths due to COVID-19 for each province and territory from 23 Apr 2020 to 6 Jan 2022. However, the testing volumes for the Yukon Territory were largely missing from 3 Jun 2021 onward. For this reason we used the testing volume data reported by Yukon Health (Government of Yukon, 2022) to supplement the PHAC data. Our third set of data was the provincial vaccination status reports, also from the PHAC (Government of Canada, 2022b). These reports include total counts per province of individuals who had received one vaccination shot, and who had received at least two vaccination shots. All count data were aggregated by week to alleviate the issue of lump sum data reporting (such as when COVID-19 cases detected over a weekend are only reported after the weekend). After aggregation, we were left with 90 weeks of data for model fitting.

#### 3.1.2 Parameter Covariates

We chose several parameter covariates for the Canada-wide model. Both *λ* and γ were allowed to vary by site (province/territory). We used weekly testing volume as a covariate for detection probability, since more testing should lead to higher detection rates. Figure S10 in the Appendix shows the testing volume data for each site. Vaccination rates (at least one dose and two doses) were used as covariates for the parameters *p*_*r*_, *p*_*d*_, *ω*_1_, *ω*_2_, and γ. Figure S11 in the Appendix shows the vaccination rates as a portion of the total population. Single dose rates are shown in red, while second dose rates are shown in blue. The vaccination rates for the Northwest Territories indicate a data anomaly in June 2021, where both single and double dose rates were reduced. This may be due to a data correction which occurred in June 2021.

The dates of emergence in Canada for the two variants of concern—Delta and Omicron—were used as covariates for time period change points for the variables γ, *ω*_1_, and *ω*_2_. As start date for the time periods, we used the first week of confirmed cases in Canada, 4 Apr 2021 (week 51) for Delta, and 28 Nov 2021 (week 84) for Omicron.

#### 3.1.3 Results

Figure 2 shows both the detected case counts *n*_*it*_ and the estimated total active cases 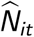 for each site (province or territory). Due to the exponential growth and decay in case numbers, the active cases are plotted on a log scale. Every site saw a large increase in cases during the Omicron time period, whereas the Delta time period saw an initial drop in cases from May to August 2021 in most sites.

**Figure 2:**
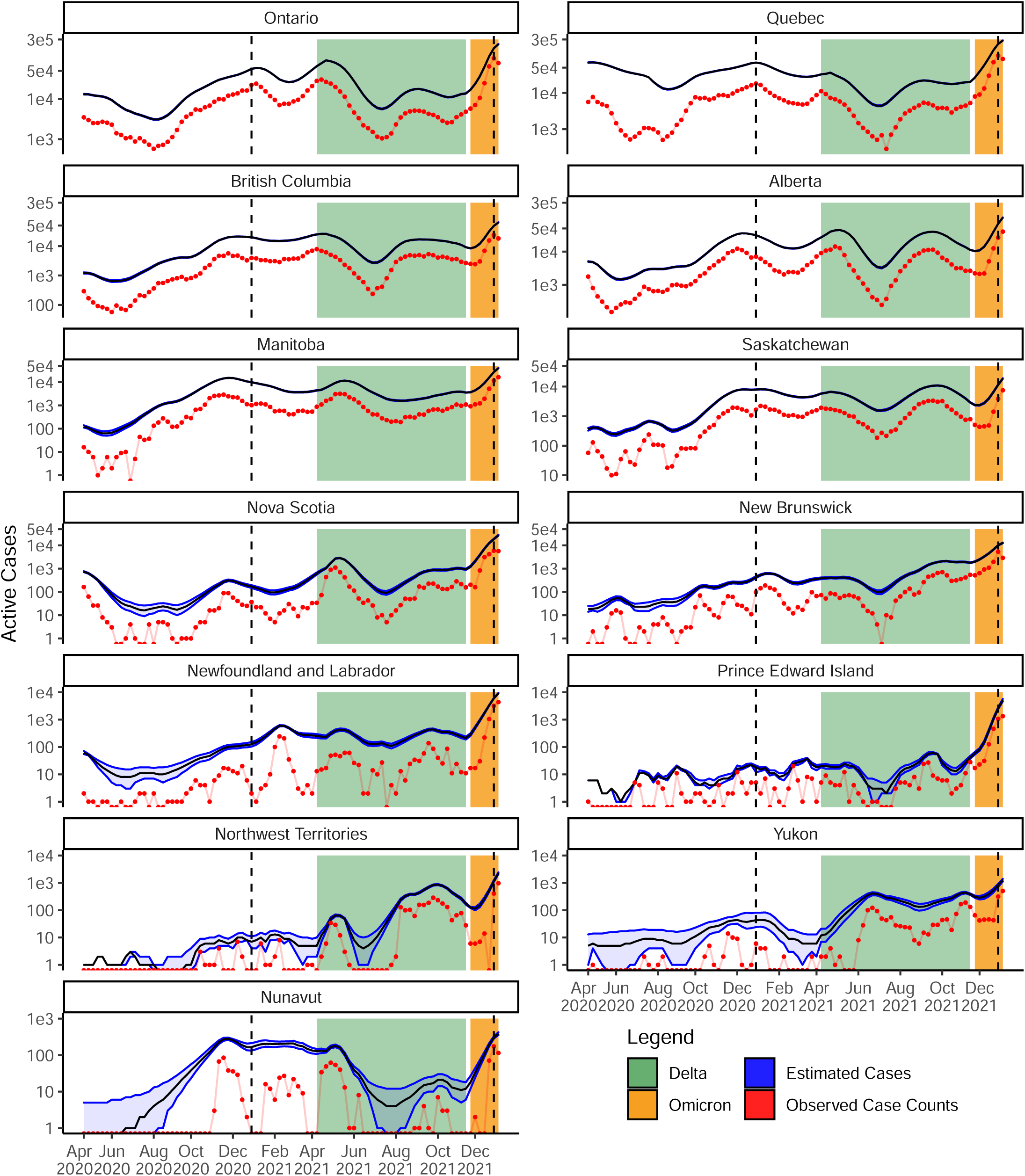
Detected case counts (red) and estimated total active cases (blue) for each province/territory from 23 Apr 2020 to 6 Jan 2022. The time periods for the two variants of concern Delta and Omicron are depicted with coloured bands. The two vertical dashed lines indicate 1 Jan 2021 and 1 Jan 2022. Active cases are plotted on a log scale. Bands indicate 95% credible intervals.

The detection probability 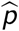 changes over time due to testing volumes (Figure 3). Detection rates in Newfoundland and Labrador, Nunavut, and Yukon were much lower than other sites for the majority of the pandemic, while the Prince Edward Island detection rate was consistently higher than 35%. Several provinces (such as B.C., Ontario, and Quebec) showed consistent growth in detection rates over the course of the pandemic, with a shared noticeable slump during the first four months of the Delta time period.

**Figure 3:**
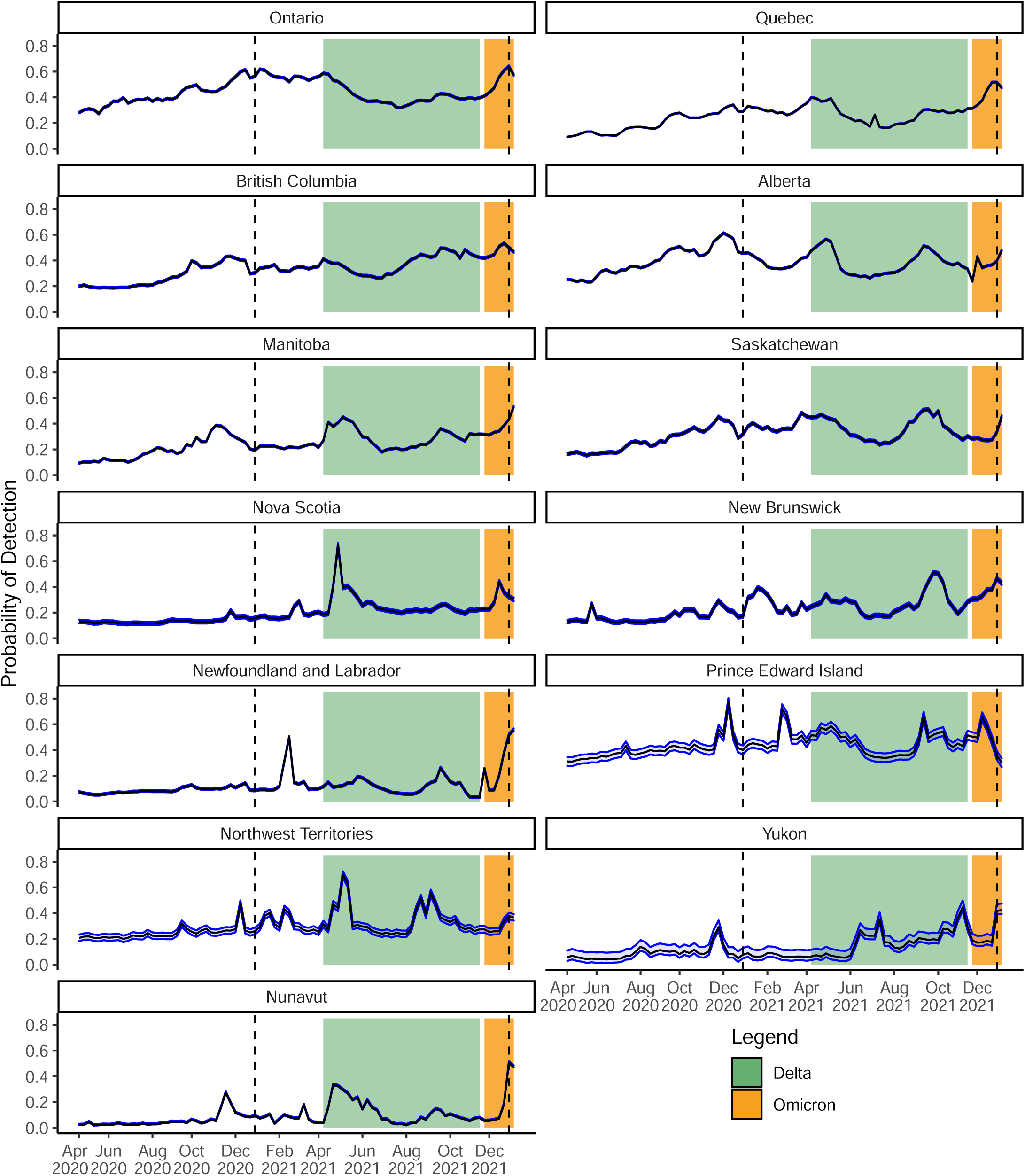
Estimated weekly detection probability (blue) for active COVID-19 cases for each province or territory from 23 Apr 2020 to 6 Jan 2022. The time periods for the two variants of concern—Delta and Omicron—are depicted with coloured bands. The two vertical dashed lines indicate 1 Jan 2021 and 1 Jan 2022. Bands show 95% credible intervals.

The probability of death, 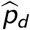, was seen to hold steady at 0.26% in all sites for the first period of the pandemic, before decreasing (Figure S12 in the Appendix). The effect of vaccinations on *p*_*d*_ is seen as a dramatic decrease throughout early 2021. The Delta period saw an increase in *p*_*d*_, which was ameliorated by increasing vaccination coverage in each site. The Omicron period saw a sharp drop in mortality, with *p*_*d*_ levelling out around 0.06% across Canada. Nunavut saw a larger probability of death during the Delta time period than the other sites, with a 95% credible interval of (0.19, 0.20)%, which does not overlap with any of the other site credible intervals. The three territories saw probability of death drop earlier than the provinces, likely due to earlier vaccination drives within the territories; Yukon reached 60% vaccination by 15 May 2021, while Saskatchewan reached 60% by 26 Jun 2021. The median death probabilities are shown in Table S1 in the Appendix.

Probability of recovery was steady at 35.0% during the early stages of the pandemic for all sites until January 2021 when the recovery probability began to increase along with the increase in vaccination coverage across Canada (Figure S13 in the Appendix). The Delta period saw a small decrease in recovery probability for all sites, while the Omicron period saw a large decrease. Probability of recovery peaked much earlier for the three territories than for any of the provinces, likely due to the earlier vaccination drives in the territories. The median recovery probabilities are shown in Table S2 in the Appendix.

The domestic spread for undetected cases, 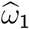, accounted for most cases in the pre-2021 period (Figure 4). As vaccination coverage increased across Canada, the spread rate for detected cases, 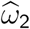, increased, and in all sites became larger than the spread rate for undetected cases during the Delta time period. During the Omicron time period, the two spread rates converged to similar levels. During the early pandemic, Prince Edward Island had the largest spread rate among undetected cases, with a 95% credible interval of (1.11, 1.26), much larger than the Canada wide average of (0.55,0.59). During this early pandemic period, Yukon had the lowest rate, with a 95% credible interval of (0.25,0.36). Conversely, the spread rate among detected cases was much smaller, with the Canada wide average having a 95% credible interval of (0.14, 0.32). In the Omicron period, the spread rate for undetected cases was less than the spread rate for detected cases, with the Canada wide 95% credible intervals of (1.17,1.21) and (1.22,1.62) respectively. The median spread rates are shown in Table S3 for undetected, and in Table S4 for detected cases in the Appendix.

**Figure 4:**
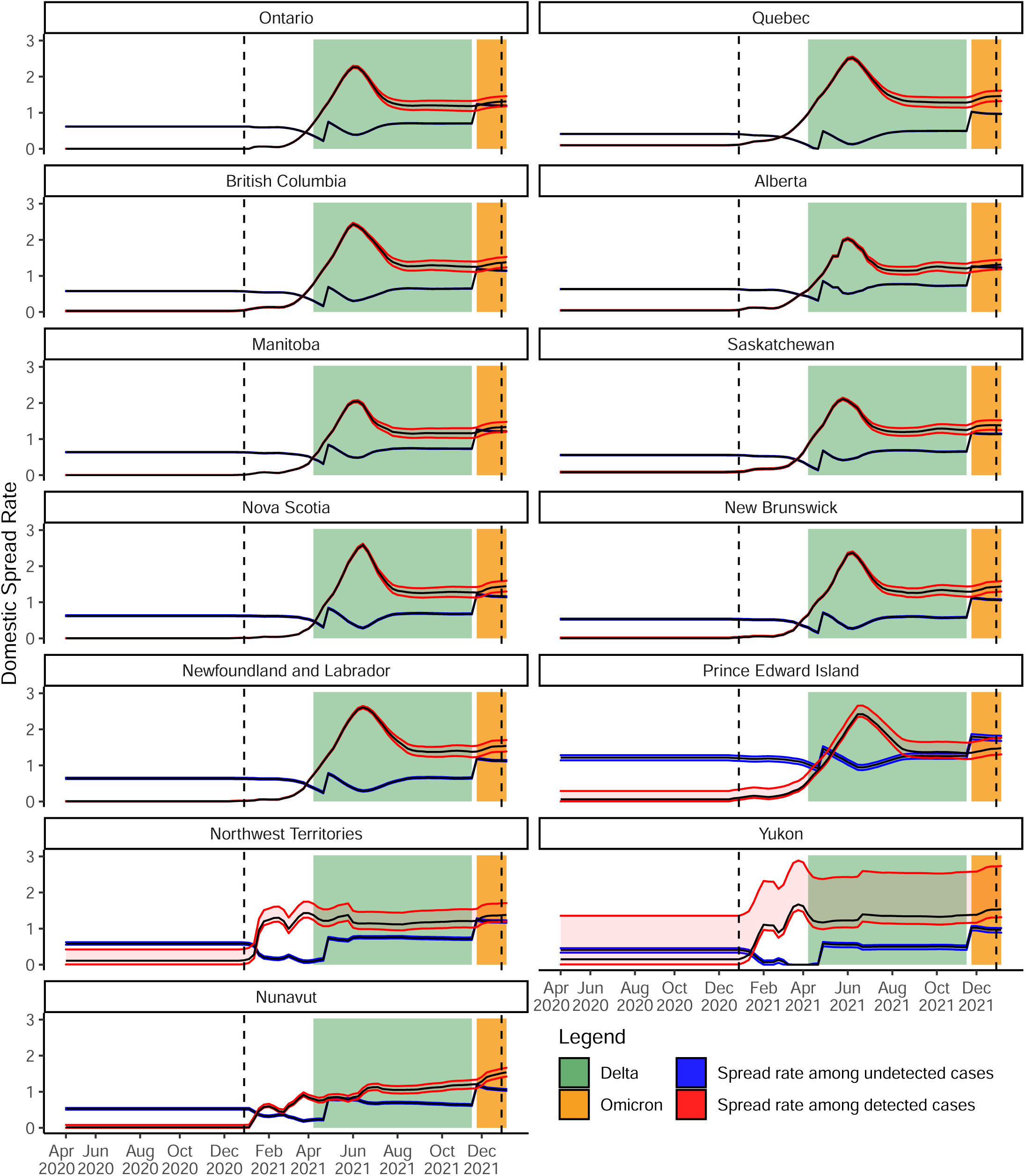
Estimated weekly domestic spread rates for detected (red, 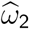) and undetected (blue, 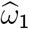) active COVID-19 cases for each province/territory from 23 Apr 2020 to 6 Jan 2022. The time periods for the two variants of concern Delta and Omicron are depicted with coloured bands. The two vertical dashed lines indicate 1 Jan 2021 and 1 Jan 2022. Bands show 95% credible intervals.

Weekly importation rates, 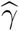, are generally low across Canada, with the exceptions of Quebec and British Columbia (Figure S14 in the Appendix). British Columbia had the second largest importation rate, at around 80 imported cases per week. Quebec had the largest, at around 1,400 imported cases per week. It is important to note that while γ is an identifiable parameter in the model, γ measures average linear growth in cases, and so is not necessarily the true importation rate. Rather, the very large γ estimate for Quebec could indicate better control of the exponential spread rate due to quarantines and other public health measures. Evidence for this can be seen in Figure 4, where the domestic spread rate among undetected cases for Quebec is much lower than for the other provinces.

To estimate the infection fatality rate (IFR) for Canada as a whole, we used the estimated new cases, 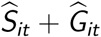 per time period *P*, then we estimated the IFR as 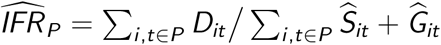. The estimated IFRs for Canada as a whole with 95% credible intervals are: 0.015 (0.012, 0.038) for the early pandemic period, 0.005 (0.004, 0.006) for the Delta period, and 0.002 (0.001, 0.003) for the Omicron period (Figure 5).

**Figure 5:**
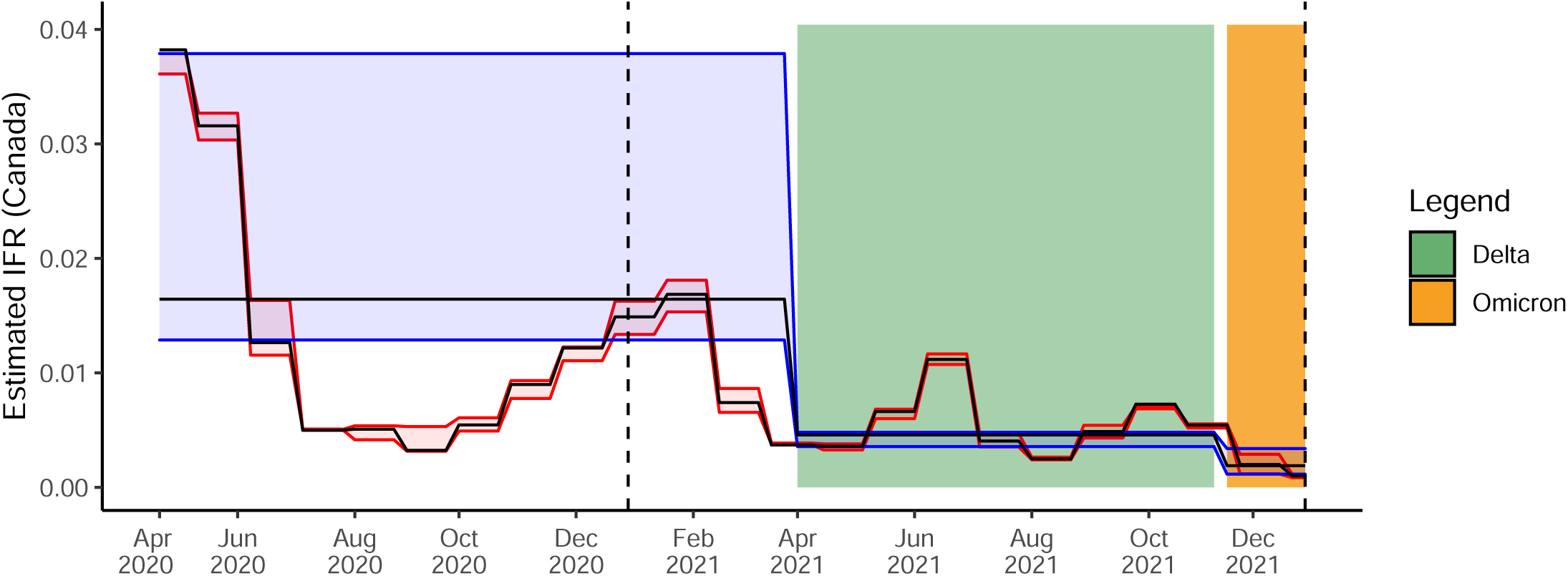
Estimated infection fatality rate (IFR) for COVID-19 infections within Canada. The time periods for the two variants of concern Delta and Omicron are depicted with coloured bands. The estimated IFR for each time period is 0.015 (early pandemic period), 0.005 (Delta period), and 0.002 (Omicron period). The two vertical dashed lines indicate 1 Jan 2021 and 1 Jan 2022. Bands show 95% credible intervals. The red shows monthly estimates, while the blue shows estimates for each of the three time periods.

### 3.2 Case Study: British Columbia

#### 3.2.1 Data Sources

We used several publicly available sources to compile the data for our B.C. case study. The B.C. Surveillance Reports (BC Centre for Disease Control, 2020) were used to gather counts of cases, recoveries, and deaths. These data are shown for each Health Authority region in Figure S8 in the Appendix. Province of B.C. laboratory data (Province of British Columbia, 2020) was used as a source for COVID-19 testing volumes per region. Start dates for the Phases of the B.C. Recovery plan were obtained from the Government of B.C. emergency preparedness response web pages (Government of British Columbia, 2020).

Data for this B.C. case study was limited to the date range 2 Apr 2020 (Week 1) to 30 Oct 2020 (Week 31). After this period, the B.C. Surveillance Reports began to exclude the data necessary for fitting these models (case counts, case recoveries, and deaths split by Health Authority region). These methods could be easily applied to more recent pandemic data if the required aggregate data were made publicly available.

#### 3.2.2 Parameter Covariates

In this case study we explored many combinations of parameter covariates in order to better understand our model, and to investigate the relationships between the covariates and the data. Health Authority Region (denoted *reg*) was used as a covariate for both *λ* and γ in all of our considered models. The covariate *reg* was also used for *ω*_1_ and *ω*_2_ in several models to indicate region dependency. Phase of the B.C. Recovery Plan (denoted *pha*) was also considered as a potential covariate for γ, *ω*_1_ and *ω*_2_, to indicate that the parameters change with time at the boundary of the recovery plan phases. For detection probability *p*, we considered the parameter covariates: *reg, pha*, COVID-19 testing volume (*vol*), and a baseline offset which was constant across regions (*B*).

#### 3.2.3 Model Selection

We used the widely applicable information criterion (WAIC: Watanabe and Opper, 2010) to aid in model selection and study the impact of covariates. Gelman et al. (2014) discuss the limitations of relying solely on log predictive density methods (such as WAIC) for model selection. In particular, the authors note that such model selection procedures can over fit the model to the data, providing suboptimal predictive performance. To that end, they caution to view information criteria as an approach to understand fitted models rather than to choose from among them.

While we do use WAIC to aid in model selection, it is not the sole criteria. For example, we select the second best performing model according to WAIC as our preferred model. We did this for three reasons: (1) the top two ranked models perform very similarly in terms of estimated latent variables, (2) the number of model parameters is far less for the second ranked model, and (3) the covariates used in the second model carry far more explanatory value than those in the first ranked model.

#### 3.2.4 Results

Our results for fitting 22 models with different organizations of the covariates and conditions on *ω* to the B.C. data are summarized in Table S5 in the Appendix. Model M1 performed best in terms of WAIC, and model M22 performed worst (we label the models in descending order of WAIC). Excluding the four models which used no covariates for *ω*_1_ and *ω*_2_, every model for which *ω*_1_ was allowed to differ from *ω*_2_ performed better than the models which forced *ω*_1_ = *ω*_2_. This is strong evidence in favour of the models which allow the domestic spread rate to be affected by detection status, since those models perform uniformly better in terms of WAIC. In addition, every model that allowed *ω*_1_ and *ω*_2_ to vary with Phase performed better than models not varying with Phase. This suggests spread rates were not constant over the course of the early pandemic.

We identify model M2 as the best model, despite M1 having the lower WAIC score. Figure S9 in the Appendix compares the estimated active cases for each Health Authority region between model M1 and model M2. The two models perform similarly for all sites, except Fraser Health Authority region, for which the model M2 predicts a substantially larger initial number of active cases. However, model M1 uses 18 more parameters than model M2, and so is at greater risk of overfitting. Model M2 uses testing volume as a covariate for detection probability, which is far more interpretable than B.C. Recovery Plan Phase (which is used instead for model M1, and is correlated with many confounding factors such as season, health mandates, and human mobility). Figure S9 also compares the results for the five single-site models (shown in green) against the two best fitted multi-site models (M1 in red and M2 in blue). The single-site models show larger uncertainty in their estimates through wide credible intervals, illustrating an advantage of using multi-site modelling.

We show the results for our chosen model M2 in Figure 6. The trend for all regions over this period of study was for a large initial number of active cases, which falls off leading up to week 10, and builds leading into the later stages of the 31 week period. All five regions showed signs of peaking at different times between weeks 17 and 26, with a second peak beginning around week 30, and likely continuing after this period of study.

**Figure 6:**
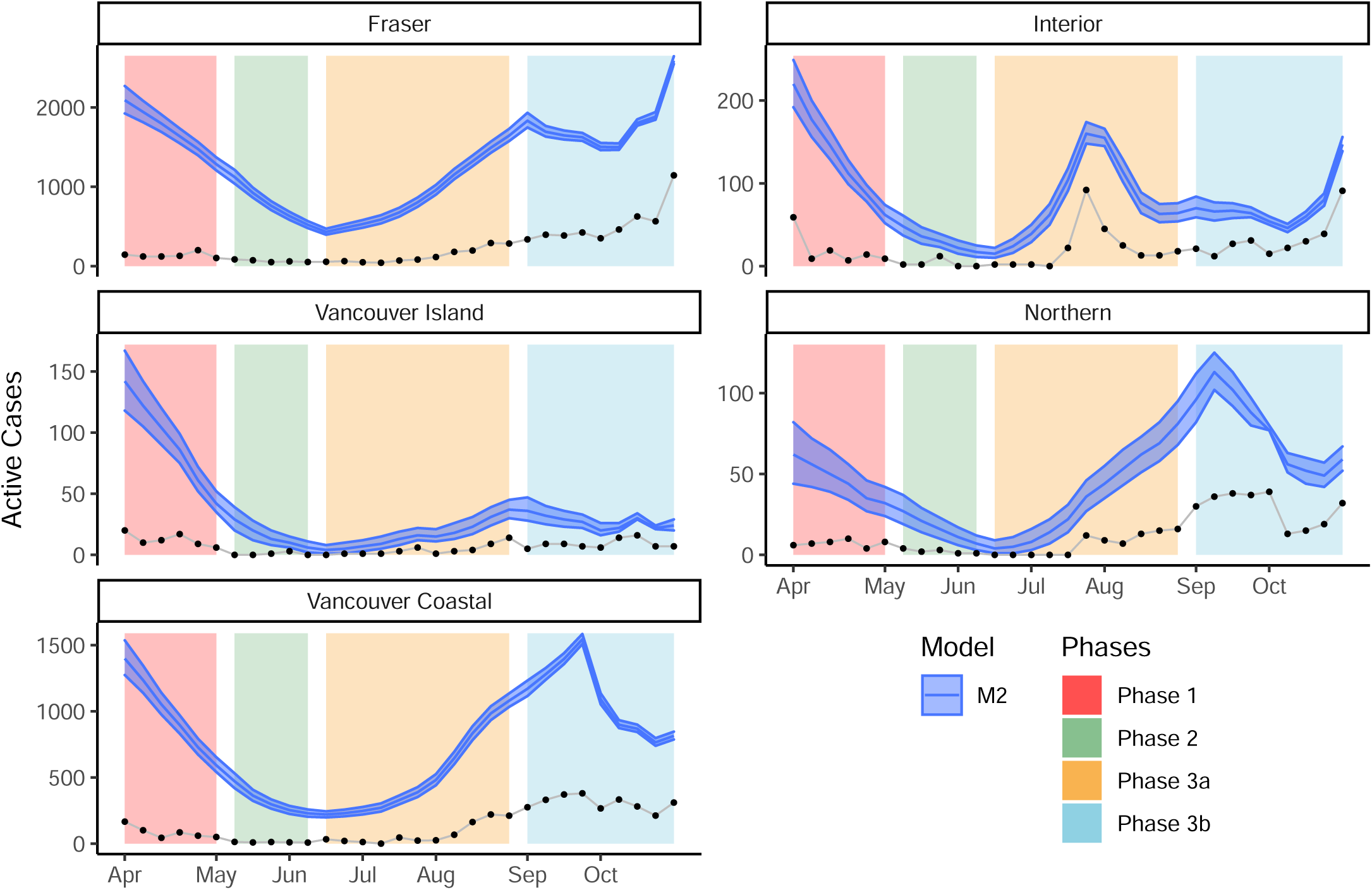
Plots of active cases split by Health Authority Region. Data starts on 2 Apr 2020 (Week 1) and ends on 30 Oct 2020 (Week 31). The blue bands show the 95% credible intervals for total active cases as estimated using model M2. The black dotted line shows the newly detected active cases each week.

### 3.3 Comparing Results

We compare results between three case studies overlapping in time intervals and regions to illustrate several important points. Model tails (time periods near the beginning and the end of the study) are less accurate, due to a lack of information outside of the study time boundaries (Section 3.3.1). This is particularly important to note when interpreting our Canada case study results. Since the Omicron time period lies on the boundary, there is less certainty in our estimates for that time period. Future applications of these methods should be aware of these boundary limitations, and should not put excessive weight on estimates near the study time boundaries. Another important point is the dependence of these models on data quality. The difference in case counts between the BCCDC Surveillance reports and the PHAC situation reports impacts the estimates of total active cases (see Figure S10 in the Appendix). However, when the data sources are the same, the multi-site models produce more precise estimates than the single-site models (Section 3.3.2). Therefore, when using these models it is important to use the most accurate and highest quality case count data available.

#### 3.3.1 British Columbia Comparison

Figure S10 in the Appendix illustrates the estimated total cases in B.C. as estimated by the single-site Health Authority Region model (green), and as estimated by the multi-site Canada case study model (blue). The two models diverge considerably in estimated total cases in both tails, while agreeing in the middle period. There are several reasons for this. First, the data used to fit the models is not the same. The detected case counts are a form of administrative data, and have been updated and corrected over time, and the counts used in the Canada model are likely to be more accurate than the counts used in the B.C. model (Figure S10). Second, the Canada model has no covariate equivalent to the B.C. Recovery Plan phases covariate of the B.C. model, thus the B.C. model allows for more granular changes in dynamics. Third, the Canada model pools information from the pandemic across Canada, allowing the province and territory estimates to be informed by national trends. Fourth, the Canada model is trained on 90 weeks of data rather than 31, and over 13 sampling sites rather than 5. The larger amount of data used for the Canada-wide model means its results are likely to be more accurate. Fifth, the B.C. model begins with three earlier weeks of data, so that the beginning tail of the model is likely more accurate than the Canada model. Conversely, the Canada model contains 62 weeks of more recent data than the B.C. model, meaning the end tail of the Canada model is likely to be more accurate.

#### 3.3.2 Northern Health Authority Comparison

We compare the Northern Health Authority Region results from the single-site model of Parker et al. (2021) with results from our multi-site model M2 (Figure 7). The single-site results were obtained using maximum likelihood methods, and so the variability shown indicates the 95% confidence interval, whereas the multi-site model uses Bayesian MCMC to obtain 95% credible intervals. The two models show excellent agreement through overlapping credible/confidence bands, and share a common maximum during week 24, but the multi-site model has increased precision over the single-site model. An important advantage to the Bayesian MCMC method of model fitting is the ability to estimate active cases even during periods when no new cases are detected, as was the case for the Northern Health region during weeks 13 through 16.

**Figure 7:**
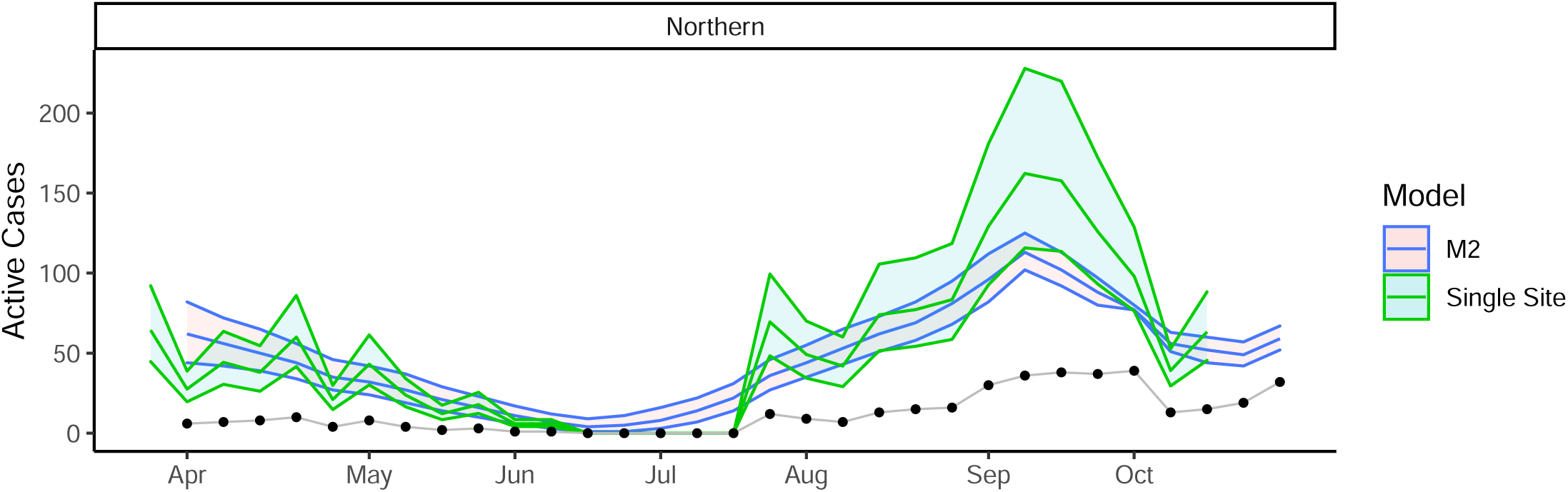
Plot of active cases for the Northern Health Authority region. Data starts on 2 Apr 2020 (Week 1) and ends on 30 Oct 2020 (Week 31). The blue band shows the 95% credible interval for total active cases as estimated using model M2. The green bands show the 95% confidence intervals for total active cases as estimated using the single site model from Parker et al. (2021), denoted Single Site. The black dotted line shows the newly detected active cases each week.

## 4 Discussion

Throughout the pandemic, case counts have been only a lower bound on the total number of active infections. Our Canada case study estimates the levels of under-reporting for each province and territory. Estimates of total active infections vary widely in magnitude across Canada and over time, with over 200,000 concurrent active cases at times in Ontario and Quebec, and under 400 concurrent active cases at any time in Nunavut. Our results show that although the first confirmed infection in Nunavut occurred in October 2020, it is likely that there were over 100 active infections in Nunavut at that time, and that the first infection likely would have occurred between June and August 2020.

Our estimates for detection probability show that there were similar detection levels across Canada. In the early pandemic, most provinces/territories had very low testing volumes (less than 10 tests per 1000 population size), which led to low detection rates (under 20% detection for most provinces/territories). Among the provinces, Prince Edward Island had the highest minimum detection rate (30%), while Newfoundland and Labrador had the lowest (3%). Among the territories, Northwest Territories had the highest minimum detection rate (20%), while Nunavut had the lowest (2%). Testing volume is not the only factor in determining detection, as can be seen by comparing the results from Ontario with those from Quebec (Figure 3). Both provinces had very similar testing volumes throughout the pandemic; however, the detection rate was substantially higher in Ontario than in Quebec. Ontario had a low of 27% and a high of 64%, compared with Quebec’s low of 10% and high of 52%. The reason for this is unclear. It could be due to differences in testing protocols, differences in access to testing, geographic or political differences, or many other possible factors. Future research into the effects of public messaging and health policies on detection rates would be beneficial and could improve our understanding of this phenomenon.

In Canada, the case-fatality rate for COVID-19 was estimated to be 4.9% in April 2020 (Abdollahi et al., 2020), and 3.36% by the end of 2020 (Shim, 2021). However, CFR is larger than IFR when there are undetected cases. Understanding the IFR during recent periods of the pandemic allows us to understand the personal risk associated with contracting SARS-CoV-2. We estimated the weekly probability of death *p*_*d*_ over the course of the pandemic, and found that for all provinces and territories of Canada the mortality rates have decreased over time. In general, mortality rates decreased with increased rate of first vaccination dose (Figure S12 in the Appendix), increased during the Delta time period, and decreased again in the Omicron time period. Weekly probability of death is different from overall probability of death, which is better measured through IFR. The weekly probability of death is the average probability of an active case dying during a particular week (if they neither die nor recover during that week, then they will have a further chance of dying in the following week and so on until they either recover or die). IFR is a more easily interpreted measure of mortality rates than *p*_*d*_, since IFR is the overall mortality rate for infected individuals. Previous findings regarding IFR using data across 15 countries determined it likely that IFR for COVID-19 was *<* 0.2% (Ioannidis et al., 2020). Fisman et al. (2020) estimated the overall IFR in Ontario to be 0.8% (0.75%, 0.85%), with a large range based on age (from 0.01% up to 12.7%). Our findings indicate that short term estimates of IFR (such as our monthly estimates in Figure 5) can fluctuate rapidly over time. Our estimate for the early pandemic time period of 1.5% is much higher than the 0.2% from Ioannidis et al. (2020). However, our monthly estimates show that depending on which month of data is considered, estimates of IFR would have varied from a high of nearly 4% down to a low of 0.3%. Our estimates for the Delta and Omicron periods are more moderate at 0.5% and 0.2%, respectively.

Estimates of recovery probability are useful for understanding the length of an average infection. During the early pandemic period the weekly probability of recovery was 35.0%, and the weekly probability of remaining an active case was 64.7%. This implies that the average recovery time and 95% recovery interval was 11.3 (6.1, 15.4) days (obtained by solving the simple geometric series in *p*_*a*_, see Appendix S.3). For the Delta and Omicron periods, the average recovery time was 8.5 (6.0, 12.8) days and 14.0 (6.1,17.0) days, respectively. We see the average recovery time increased during the Omicron period, while the lower bound on recovery time has remained around 6 days throughout the pandemic.

According to the National Collaborating Centre for Infectious Diseases (2022), both the Delta and Omicron variants of concern have been found to be more transmissible than the earlier variants. Thus we would expect to find an increased *ω*_1_ and *ω*_2_ during those time periods. Our Canada model agrees with this expectation (Figure 4), where domestic spread rates are seen to increase across Canada at the Delta boundaries as well as at the Omicron boundaries. A single vaccine dose is correlated with decreased spread rate for the undetected cases (corresponding loosely to asymptomatic, pre-symptomatic, and low severity cases), and is correlated with increased spread rate for the detected cases (corresponding loosely to symptomatic and medium to high severity cases). The increased spread rate could be explained by decreases in severity for vaccinated individuals (Lauring et al., 2022), so that the detected cases are more inclined to mobility and interaction events, as well as relaxed health measures correlated with increasing vaccination rates. Receiving a second vaccine dose is correlated with an increase in spread rate for undetected cases, which may be explained by a decrease in perceived personal danger from contracting the disease when vaccinated with two doses. The second vaccine dose is correlated with a large decrease in spread rate for the detected cases in all provinces, but not in the territories.

Our British Columbia case study was limited in scope by a lack of available public data after 30 Oct 2020. For future pandemics and disease outbreaks, we would urge all public health authorities (in B.C., in Canada, and abroad) to make weekly aggregate counts of cases, recoveries, and deaths publicly accessible as early as possible to promote greater knowledge and more expedient research. The decision to do so can in turn lead to better informed, more timely health policy decision making, which can save lives and reduce the burden on our health care systems.

## Data Availability

All data are available online at: https://github.com/mrparker909/COVID-MultiSiteModel2022.git

## Acknowledgments

LC was supported by a Michael Smith Foundation for Health Research and Victoria Hospitals Foundation Grant #COV-2020-1061 and a Canadian Statistical Sciences Institute Rapid Response Program: COVID-19.

## Supplementary Material

### S.1 Single-Site Model

Here we describe the mathematical structure of the single-site model. We drop the site subscript *i* from the multi-site model to produce the single-site model. Our model makes use of three sets of reported data: The observed case counts {*n*_*t*_}, the recoveries among case counts {*r*_*t*_}, and the deaths due to the disease {*D*_*t*_}. This description mirrors the multi-site model from Section 2.1, but with some simplifications due to the dropped subscript.

1. Initial Abundance: *N*_1_ ∼ Poisson(*λ*)
2. State Process: {*A*_*t*_, *D*_*t*_, *R*_*t*_} ∼ Multinomial(*N*_*t*_; *p*_*a*_, *p*_*d*_, *p*_*r*_)
3. Observed Active Cases: *a*_*t*_ = *n*_*t*_ + *a*_*t*−1_ − *r*_*t*−1_ − *d*_*t*−1_, *a*_0_ = 0
4. Domestic Spread: *S*_*t*_ ∼ Poisson(Ω_*t*−1_), for *t* > 1
5. Ω_*t*−1_ : *ω*_1_(*N*_*t*−1_ − *a*_*t*−1_) · *δ* + *ω*_2_*a*_*t*−1_
6. *δ* : (*H* − *N*_*t*_) *H*, where *H* is total population size
7. Imported Cases: *G*_*t*_ ∼ Poisson(γ), for *t* > 1
8. Abundance Updates: *N*_*t*_ = *A*_*t*−1_ + *S*_*t*_ + *G*_*t*_, for *t* > 1
9. Observation Process I: *n*_*t*_ ∼ Binomial(*N*_*t*_ − *a*_*t*−1_, *p*)
10. Observation Process II: 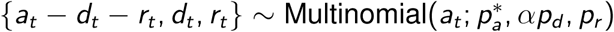

1. The initial abundance. *N*_*t*_ is the total (unknown) number of active cases at time *t. N*_1_ is the initial number of active cases at time *t* = 1. The parameter *λ* is the expected initial number of cases.
2. The multinomial state process. Each active case in *N*_*t*_ is partitioned at time *t* into one of three categories according to whether the case will remain active, die, or recover by time *t* + 1. *A*_*t*_ is the subset of *N*_*t*_ which remains active (with probability *p*_*a*_). *D*_*t*_ is the subset of *N*_*t*_ which will die (with probability *p*_*d*_). *R*_*t*_ is the subset of *N*_*t*_ which will recover (with probability *p*_*r*_). The parameter *p*_*a*_ is determined, since *p*_*a*_ = 1 − *p*_*r*_ − *p*_*d*_.
3. The observed active cases *a*_*t*_ is a deterministic quantity, specified by the observed data. It is the number of observed cases which are still active at time *t. a*_*t*_ is calculated recursively, with *a*_0_, *r*_0_, *d*_0_, *n*_0_ all zero by definition, unless observed active cases are known for the previous time period, in which case *a*_0_ would not be zero. Note that if deaths were fully observed, *d*_*t*_ would be equal to *D*_*t*_.
4. *S*_*t*_ models the domestic spread, which is the spread of the disease due to contact with infectious individuals within the population. Ω_*t*_ is the average number of new infections per time interval, which is calculated using the two parameters *ω*_1_ and *ω*_2_. *S*_*t*_ allows for exponential growth of cases, but does not allow for a spontaneous outbreak within a population that has zero active cases.
5. Ω_*t*_ has a similar meaning to the reproductive number *R*_0_. However, unless the time interval is the same as the infectious period, Ω_*t*_ is in general different from *R*_0_. The parameter *ω*_1_ is the average new infections per unobserved active case *N*_*t*−1_ − *a*_*t*−1_. The parameter *ω*_2_ is the average number of new infections per observed active case *a*_*t*−1_.
6. *δ* is a moderating term which modulates the growth of cases as the population becomes saturated with cases. *H* is the total population size of the region, which is the maximum number of infected individuals that are possible. *δ* decreases to zero linearly as *N*_*t*_ approaches *H*. Thus, the spread rate goes to zero as the population becomes saturated with cases (we assume that the spread rate is proportional to the number of susceptible individuals).
7. *G*_*t*_ models the number of imported cases, which are new cases entering the population (from travel for example). The parameter γ is the average number of new imported cases per time interval. Imported cases allow for disease to occur even when *N*_1_ = 0. This can allow for spontaneous outbreaks in regions with no previous active cases. *G*_*t*_ allows for linear growth of cases over time.
8. The abundance updates. Because we assume that the number of cases changes with time, we calculate the abundance after time 1 using *A*_*t*_ from the state process in addition to the growth terms *S*_*t*_ and *G*_*t*_. In this way, *N*_*t*_ are previous cases which have remained active (*A*_*t*−1_) plus the new cases due to domestic spread (*S*_*t*_) and the new cases due to importation (*G*_*t*_).
9. The first observation process models the reporting of case counts. The parameter *p* is the probability of detecting a case, and so 1 − *p* gives the under-reporting rate. The usual N-mixture model would have *n*_*t*_ ∼ Binomial(*N*_*t*_, *p*). However, since *N*_*t*_ is the total number of active cases, this would allow double counting (because observed cases *n*_*t*_ are tracked until recovery or death). Instead, we subtract the already observed active cases prior to the binomial thinning: *N*_*t*_ − *a*_*t*−1_.
10. The second observation process models reporting of recoveries and deaths. Similar to the state process for *N*_*t*_, the states *a*_*t*_ are partitioned into cases that remain active, cases that die, and cases that recover. If deaths are assumed fully detected, *α* = 1*/p* would account for perfect detection of deaths (which increases the proportion of observed deaths among detected cases unless *p* = 1), and we would set *d*_*t*_ to *D*_*t*_ where appropriate. In the situation where deaths are under-reported, we would set *α* = 1. The parameter 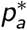 is determined for the same reason that *p*_*a*_ is determined.

The single-site model contains seven estimable model parameters: *λ, p, p*_*r*_, *p*_*d*_, γ, *ω*_1_, and *ω*_2_. It contains three sets of observed data: {*n*_*t*_}, {*d*_*t*_}, {*r*_*t*_}. And it contains six sets of latent variables: {*N*_*t*_}, {*D*_*t*_}, {*R*_*t*_}, {*A*_*t*_}, {*S*_*t*_}, {*G*_*t*_}. In the situation where deaths are fully observed, such as in our two case studies, {*D*_*t*_} is no longer a latent variable, and replaces {*d*_*t*_} as observed data. This model is an extension and major update to the single-site model from Parker et al. (2021).

### S.2 Simulations

We conduct simulations to investigate the identifiability of all parameters. This is particularly important for the parameter *ω*_1_ which is only implicity tied to the data {*n*_*it*_} through the unobserved quantities {*N*_*it*_}. We set the number of sampling sites to be 2 and the number of sampling occasions to be 30. We chose one set of ground truth parameter values for this simulation: *λ* = 50, γ = 5, *ω*_1_ = 0.25, *ω*_2_ = 0.5, *p*_*r*_ = 0.4, *p*_*d*_ = 0.1, *p* = 0.7.

The parameter values for the simulation were chosen to be of reasonable magnitude, and to avoid parameter boundaries. We chose *λ* to be 50 to avoid the slow pandemic start situation of *λ* = 0, which could cause the initial few time steps to have no new cases except through importation, until enough cases existed for domestic spread to take hold. We chose γ to be an appreciable fraction of *λ* so that linear growth would be immediately impactful to the growth of case counts. A smaller γ would likely incur larger variance in the posterior distributions for γ. We chose to have *ω*_1_ and *ω*_2_ set between 0 and 1 to produce moderate spread rates, and we chose for them to be different from each other in order to test their identifiability. We chose *p*_*r*_ to be 0.4 as that was expected to be close to the actual recovery probability for Canada. We chose *p*_*d*_ to be 0.1 so that the number of deaths would be small but appreciable for the simulation study. The choice of *p* = 0.7 was arbitrary, and away from zero or 1 (which may be corner cases and which are unrealistic). In particular *p* = 1 would imply that every case was identified at all times. This is unreasonable even if a population census was performed at each time step, since it would also require perfect testing results. Similarly, *p* = 0 would imply that all case counts were zero for every time step, which would mean there were no data available at all. The value *p* = 0.7 was chosen as a reasonable guess as to the maximum probability of detection across Canada.

Our simulation consisted of 50 runs. For each run a random set of population data ({*n*_*it*_}, {*r*_*it*_}, {*D*_*it*_}) was generated using the model definition. The data generating algorithm is shown in Listing S1. We chose to use two sampling sites and thirty sampling occasions for the simulation. The model was then fit to the generated data using Bayesian MCMC with a burnin of 100,000, thinning of 200, for a total of 4,500 iterations after burn-in and thinning. The resulting posterior means were collected for each parameter. The simulation results are shown in Figure S8, along with the prior distributions chosen for each parameter. Each parameter was found to be identifiable (Figure S8). The parameter with the least degree of certainty (as indicated by the wide distribution of the posterior means) was *p*, showing that the probability of detection requires more data to estimate accurately than the other model parameters. In particular, *p*_*r*_ and *p*_*d*_ are estimated with very high accuracy and precision. The two domestic spread parameters *ω*_1_ and *ω*_2_ are estimated with higher precision than the importation rate γ, but with slightly less accuracy. We expect that the parameter estimate accuracy and precision would increase for all parameters with increasing sampling occasions *T*, sampling sites *M*, and simulation runs.

**Listing S1:**
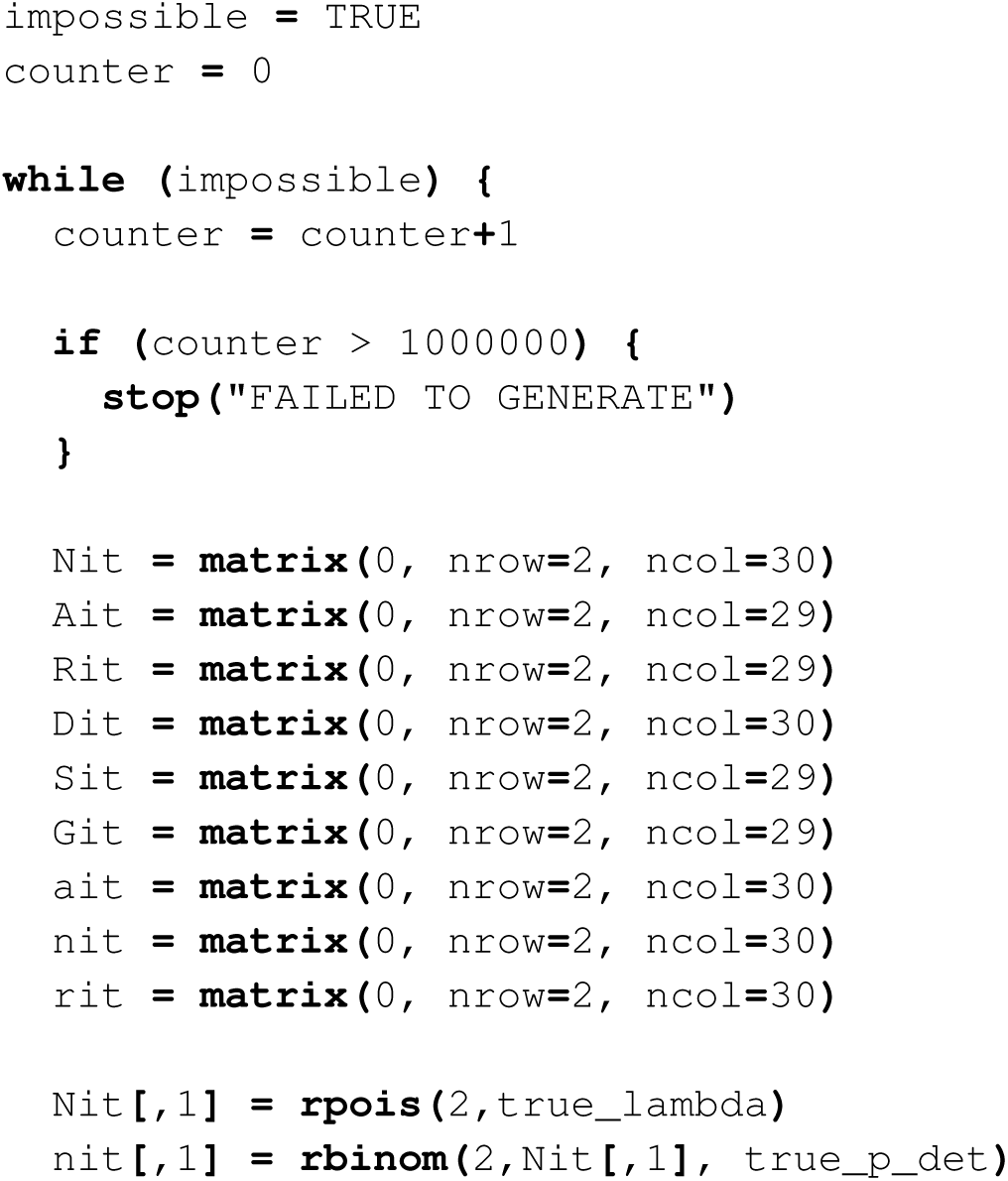
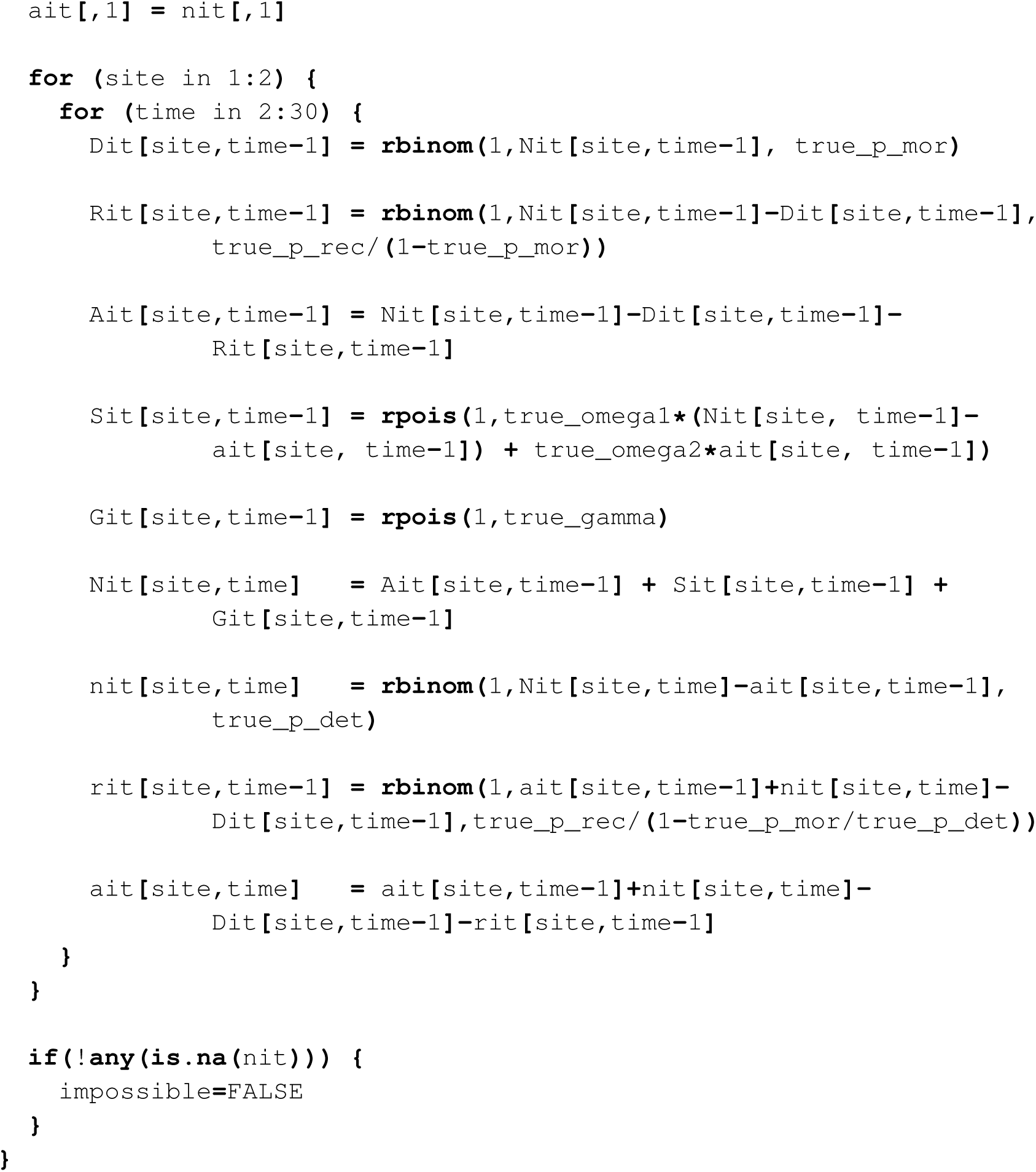
R code for data generating algorithm for simulation study.

Figure S9 shows the running posterior means for each parameter for one run of the simulation (run 25 out of 50). The posterior means are each seen to converge close to the ground truth values rapidly, with only small changes after 1,000 iterations. The stability of the running posterior means shows good mixing and convergence of the posterior distributions. The other simulation runs had similar results.

For N-mixture type models, there has been substantial literature discussing lack of identifiability for *p* when the counts {*n*_*it*_} are small and when the number of sampling occasions *T* is small (Dennis et al., 2015; Barker et al., 2018). Our model may avoid this small *p* problem entirely due to the use of additional data through {*r*_*it*_} and {*D*_*it*_}, as well as the additional model structure through the multinomial state process. However, should this work be applied to models fit with very small count sizes and very few sampling occasions, additional simulation studies may be required to verify parameter identifiability in that setting.

### S.3 Average Recovery Times

Calculating average recovery time 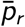 using weekly probability of recovery (*p*_*r*_) and weekly probability of remaining an active case (*p*_*a*_) requires solving a simple geometric series in *p*_*a*_. The geometric series arises from the repeated chance of recovery for each week of remaining active, and can be seen as the infinite sum: 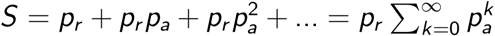. We can consider the partial sums 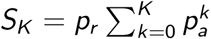. Then, *S*_*K*_ is the total probability of recovery up to week *K*.

When *S*_*K*_ = 0.5, *K* + 1 is the number of weeks required for an average recovery. Note that it is *K* + 1 and not *K* due to the definition of *p*_*r*_ and *p*_*a*_ as probabilities between sampling occasions. The well known partial sum solution for the geometric series is: 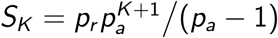. Rearranging to solve for *K* :

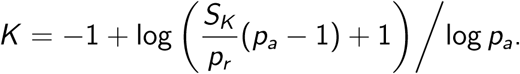

Using our Canada wide estimates for the early pandemic time period (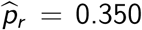, and 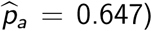), we calculate *K* + 1 = 1.61. This means that the expected recovery time is, 161% of one week, or 11.3 days 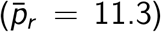. Our reported 95% recovery interval of (6.1, 15.4) days for the early pandemic period was calculated using the exact same procedure but with *S*_*K*_ = 0.025 for the lower bound and *S*_*K*_ = 0.975 for the upper bound.

### S.4 Supplementary Tables

**Table S1:**
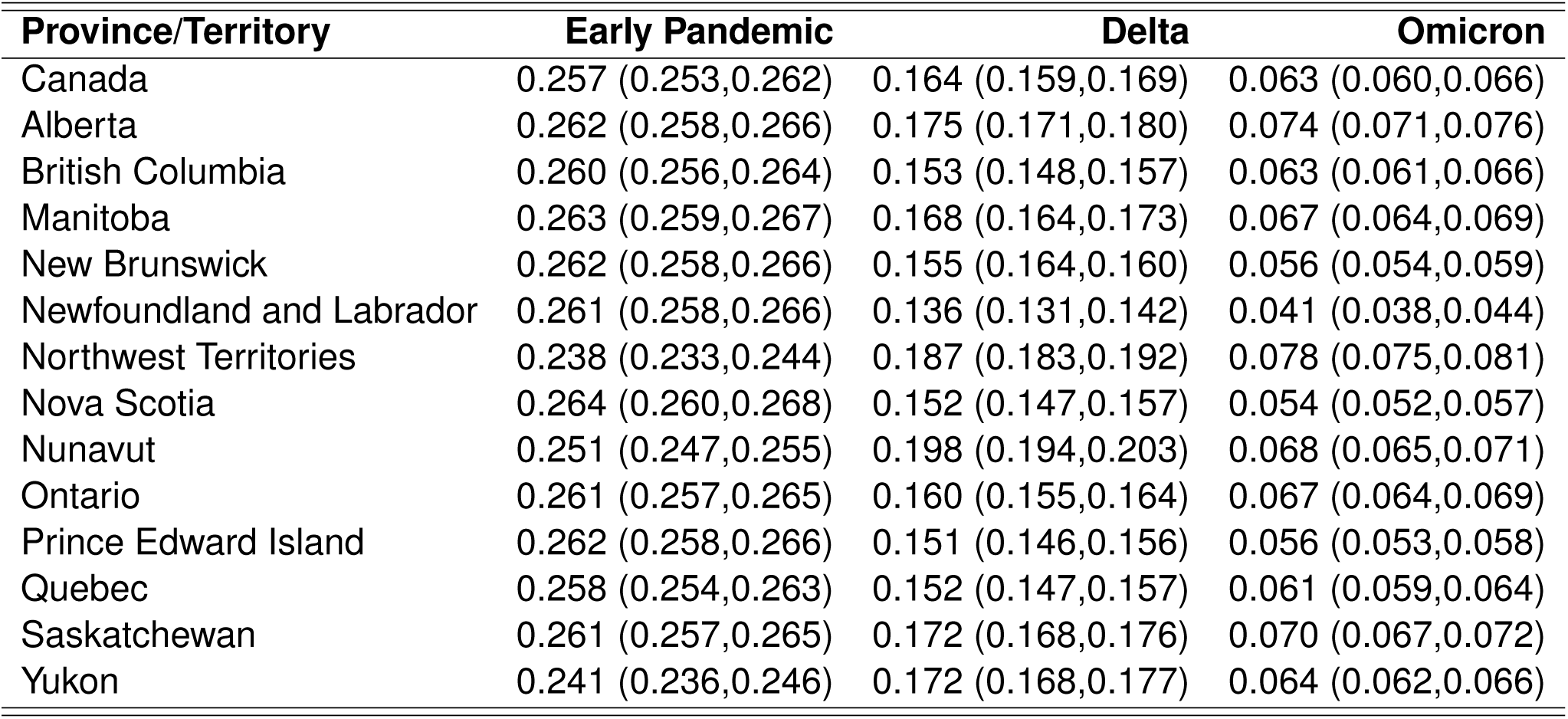
Average weekly probabilities of death in percent (%) for active COVID-19 cases (including unreported cases) split by three periods of the pandemic: Early Pandemic (prior to 4 Apr 2021), Delta (between 4 Apr and 28 Nov 2021), and Omicron (post 28 Nov 2021). 95% credible intervals are shown in parentheses.

**Table S2:** Average weekly probabilities of recovery in percent (%) for active COVID-19 cases split by three periods of the pandemic: Early Pandemic (prior to 4 Apr 2021), Delta (between 4 Apr and 28 Nov 2021), and Omicron (post 28 Nov 2021). 95% credible intervals are shown in parentheses.

| Province/Territory | Early Pandemic | Delta | Omicron |
| --- | --- | --- | --- |
| Canada | 35.0 (34.9,35.0) | 43.4 (43.3,43.5) | 29.3 (29.2,29.4) |
| Alberta | 34.5 (34.5,34.6) | 42.5 (42.4,42.6) | 28.4 (28.4,28.5) |
| British Columbia | 34.7 (34.7,34.8) | 45.0 (44.9,45.1) | 29.0 (28.9,29.0) |
| Manitoba | 34.4 (34.3,34.4) | 43.0 (42.8,43.1) | 28.9 (28.8,29.0) |
| New Brunswick | 34.5 (34.5,34.6) | 44.7 (44.6,44.8) | 29.8 (29.7,29.9) |
| Newfoundland and Labrador | 34.6 (34.5,34.7) | 47.0 (46.9,47.2) | 30.8 (30.8,30.9) |
| Northwest Territories | 36.8 (36.7,36.9) | 39.8 (39.7,39.9) | 28.3 (28.2,28.4) |
| Nova Scotia | 34.3 (34.2,34.3) | 45.0 (44.9,45.1) | 29.8 (29.7,29.9) |
| Nunavut | 35.5 (35.4,35.5) | 39.5 (39.4,39.6) | 30.2 (30.2,30.3) |
| Ontario | 34.6 (34.5,34.7) | 44.1 (44.0,44.2) | 28.7 (28.7,28.8) |
| Prince Edward Island | 34.4 (34.4,34.5) | 45.3 (45.2,45.4) | 29.5 (29.5,29.6) |
| Quebec | 35.0 (34.9,35.1) | 45.0 (44.9,45.1) | 29.2 (29.1,29.3) |
| Saskatchewan | 34.7 (34.6,34.7) | 43.0 (42.9,43.1) | 28.9 (28.9,29.0) |
| Yukon | 36.5 (36.4,36.6) | 40.6 (40.5,40.7) | 29.1 (29.1,29.2) |

**Table S3:** Average weekly domestic spread rates for unobserved cases (*ω*_1_) split by three periods of the pandemic: Early Pandemic (prior to 4 Apr 2021), Delta (between 4 Apr and 28 Nov 2021), and Omicron (post 28 Nov 2021). 95% credible intervals are shown in parentheses.

| Province/Territory | Early Pandemic | Delta | Omicron |
| --- | --- | --- | --- |
| Canada | 0.57 (0.55,0.59) | 0.65 (0.63,0.67) | 1.19 (1.17,1.21) |
| Alberta | 0.61 (0.60,0.61) | 0.71 (0.70,0.72) | 1.25 (1.24,1.25) |
| British Columbia | 0.54 (0.54,0.55) | 0.57 (0.56,0.57) | 1.16 (1.16,1.17) |
| Manitoba | 0.61 (0.61,0.62) | 0.69 (0.68,0.70) | 1.23 (1.22,1.24) |
| New Brunswick | 0.50 (0.49,0.52) | 0.53 (0.52,0.55) | 1.09 (1.07,1.10) |
| Newfoundland and Labrador | 0.61 (0.59,0.63) | 0.56 (0.54,0.58) | 1.15 (1.13,1.17) |
| Northwest Territories | 0.47 (0.44,0.51) | 0.74 (0.70,0.77) | 1.21 (1.18,1.25) |
| Nova Scotia | 0.61 (0.59,0.62) | 0.61 (0.60,0.63) | 1.18 (1.16,1.19) |
| Nunavut | 0.46 (0.44,0.49) | 0.70 (0.68,0.73) | 1.09 (1.07,1.12) |
| Ontario | 0.59 (0.58,0.59) | 0.63 (0.63,0.64) | 1.21 (1.20,1.22) |
| Prince Edward Island | 1.19 (1.11,1.26) | 1.19 (1.12,1.26) | 1.77 (1.70,1.84) |
| Quebec | 0.37 (0.37,0.37) | 0.40 (0.39,0.40) | 0.99 (0.98,0.99) |
| Saskatchewan | 0.52 (0.51,0.53) | 0.62 (0.61,0.63) | 1.15 (1.14,1.16) |
| Yukon | 0.32 (0.25,0.36) | 0.52 (0.44,0.56) | 0.99 (0.91,1.04) |

**Table S4:** Average weekly domestic spread rates for observed cases (*ω*_2_) split by three periods of the pandemic: Early Pandemic (prior to 4 Apr 2021), Delta (between 4 Apr and 28 Nov 2021), and Omicron (post 28 Nov 2021). 95% credible intervals are shown in parentheses.

| **Province/Territory** | **Early Pandemic** | **Delta** | **Omicron** |
| --- | --- | --- | --- |
| Canada | 0.17 (0.14,0.32) | 1.45 (1.34,1.66) | 1.37 (1.22,1.62) |
| Alberta | 0.12 (0.12,0.13) | 1.35 (1.27,1.44) | 1.28 (1.15,1.42) |
| British Columbia | 0.13 (0.12,0.14) | 1.57 (1.48,1.67) | 1.32 (1.18,1.47) |
| Manitoba | 0.07 (0.07,0.07) | 1.35 (1.26,1.45) | 1.28 (1.15,1.43) |
| New Brunswick | 0.09 (0.08,0.11) | 1.52 (1.43,1.62) | 1.38 (1.24,1.53) |
| Newfoundland and Labrador | 0.09 (0.09,0.10) | 1.74 (1.65,1.84) | 1.49 (1.33,1.65) |
| Northwest Territories | 0.41 (0.30,0.73) | 1.19 (1.02,1.51) | 1.34 (1.16,1.66) |
| Nova Scotia | 0.06 (0.05,0.06) | 1.55 (1.46,1.64) | 1.38 (1.23,1.53) |
| Nunavut | 0.19 (0.17,0.26) | 1.05 (0.95,1.15) | 1.41 (1.30,1.54) |
| Ontario | 0.09 (0.09,0.09) | 1.46 (1.37,1.55) | 1.27 (1.13,1.42) |
| Prince Edward Island | 0.13 (0.08,0.37) | 1.64 (1.52,1.90) | 1.42 (1.25,1.71) |
| Quebec | 0.22 (0.22,0.23) | 1.64 (1.55,1.74) | 1.41 (1.27,1.56) |
| Saskatchewan | 0.18 (0.16,0.19) | 1.43 (1.35,1.52) | 1.37 (1.24,1.50) |
| Yukon | 0.43 (0.28,1.63) | 1.31 (1.11,2.51) | 1.47 (1.25,2.67) |

**Table S5:** Results from fitting our multi-site model with various covariate structures. All models used site as a covariate for *λ*. All models considered no covariates for *p*_*d*_ and *p*_*r*_. Each parameter column indicates which covariates were considered for the given parameter. Phase of B.C. Recovery Plan is indicated by *pha*. Health Authority Region is indicated by *reg*. Virus testing volume is indicated by *vol*. A baseline offset which is the same for each region is indicated by *B*. Models for which *ω*_1_ = *ω*_2_ are indicated as such in the *ω*_2_ column (if *ω*_1_ = *ω*_2_ is not indicated here, *ω*_1_ and *ω*_2_ are not forced to be equal). Column **Q** shows the number of trained parameters in each model. Column **WAIC** shows the WAIC for each model, while Δ**WAIC** shows the difference in WAIC compared with the best performing model.

| Model | $\gamma$ | $\omega_1$ | $\omega_2$ | $p$ | Q | WAIC | ΔWAIC |
| --- | --- | --- | --- | --- | --- | --- | --- |
| M1 | (*pha, reg*) | (*pha, reg*) | (*pha, reg*) | (*pha, reg*) | 43 | 5674 | 0 |
| M2 | (*pha, reg*) | (*pha*) | (*pha*) | (*vol*) | 25 | 6223 | 549 |
| M3 | (*pha, reg*) | (*pha, reg*) | (*pha, reg*) | (*vol*) | 35 | 6234 | 560 |
| M4 | (*pha, reg*) | (*pha, reg*) | (*pha, reg*) | (*pha, vol*) | 39 | 6257 | 583 |
| M5 | (*pha, reg*) | (*pha*) | (*pha*) | (*pha, reg*) | 33 | 6316 | 642 |
| M6 | (*pha, reg*) | (*pha*) | (*pha*) | (*B, vol*) | 26 | 6332 | 658 |
| M7 | (*pha, reg*) | (*pha, reg*) | (*pha, reg*) | (*pha, reg, vol*) | 44 | 6337 | 663 |
| M8 | (*pha, reg*) | (*pha, reg*) | (*pha, reg*) | (*reg, vol*) | 40 | 6442 | 768 |
| M9 | (*pha, reg*) | (*pha*) | (*pha*) | (*reg, vol*) | 30 | 6454 | 780 |
| M10 | (*pha, reg*) | (*pha, reg*) | (*pha, reg*) | (*B, vol*) | 36 | 6517 | 843 |
| M11 | (*pha, reg*) | (*pha, reg*) | $\omega_1 = \omega_2$ | (*pha, reg, vol*) | 35 | 6541 | 867 |
| M12 | (*pha, reg*) | (*pha*) | $\omega_1 = \omega_2$ | (*vol*) | 21 | 6625 | 951 |
| M13 | (*pha, reg*) | (*pha, reg*) | $\omega_1 = \omega_2$ | (*pha, vol*) | 30 | 6658 | 984 |
| M14 | (*pha, reg*) | (*pha*) | $\omega_1 = \omega_2$ | (*pha, reg*) | 29 | 6695 | 1021 |
| M15 | (*pha, reg*) | (*pha*) | $\omega_1 = \omega_2$ | (*pha, reg, vol*) | 30 | 6702 | 1028 |
| M16 | (*pha, reg*) | (*pha*) | $\omega_1 = \omega_2$ | (*pha, vol*) | 25 | 6773 | 1099 |
| M17 | (*pha, reg*) | (*pha*) | $\omega_1 = \omega_2$ | (*reg, vol*) | 26 | 6798 | 1124 |
| M18 | (*pha, reg*) | (*pha*) | $\omega_1 = \omega_2$ | (*B, vol*) | 22 | 6868 | 1194 |
| M19 | (*reg*) | (none) | (none) | (*reg, vol*) | 20 | 7039 | 1365 |
| M20 | (*reg*) | (none) | (none) | (*B, vol*) | 16 | 7058 | 1384 |
| M21 | (*reg*) | (none) | (none) | (*pha, reg*) | 23 | 7131 | 1384 |
| M22 | (*reg*) | (none) | (none) | (*pha, reg, vol*) | 24 | 7304 | 1630 |

### S.5 Supplementary Figures

**Figure S8:**
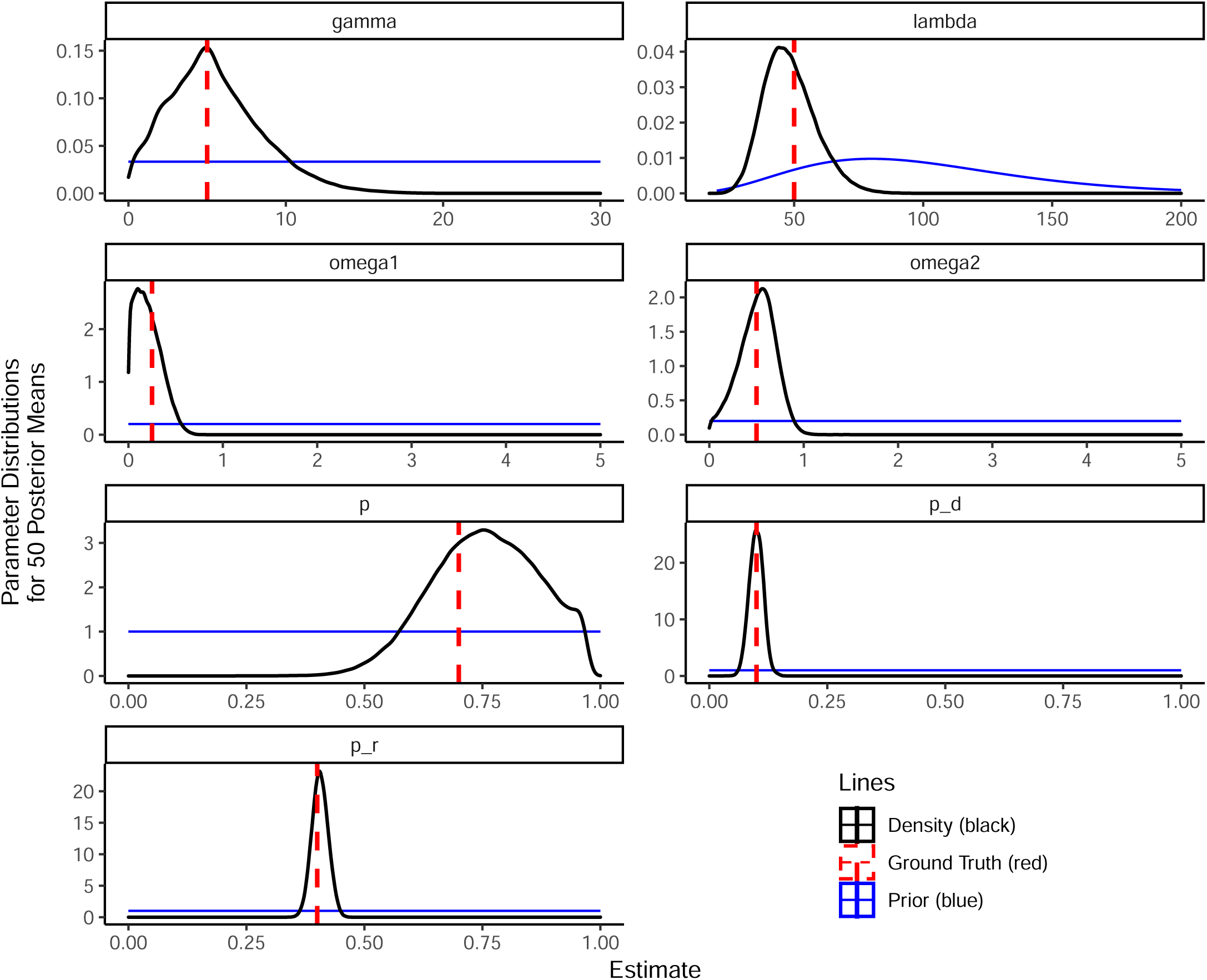
Density plots showing posterior means for each estimated parameter. Fifty simulation runs were used. Red dashed lines indicate the ground truth parameter values for the simulation. Posterior modes are close to ground truth parameter values.

**Figure S9:**
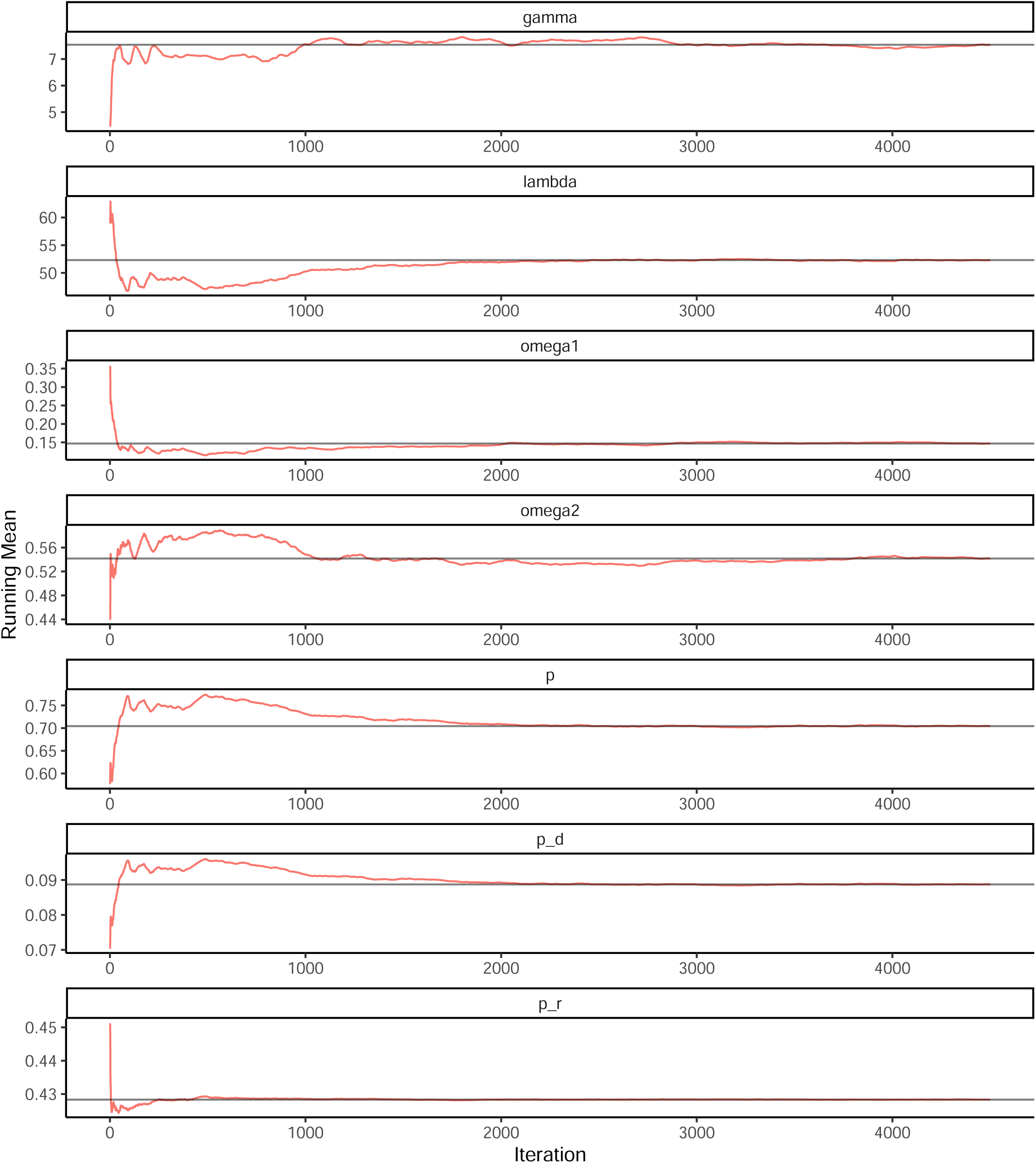
Posterior running means for each estimated parameter for a single fixed replicate (dataset 25 out of 50) of the simulation study. The black horizontal lines indicate the overall mean. Red lines indicate the running means. Running means approach overall means.

**Figure S10:**
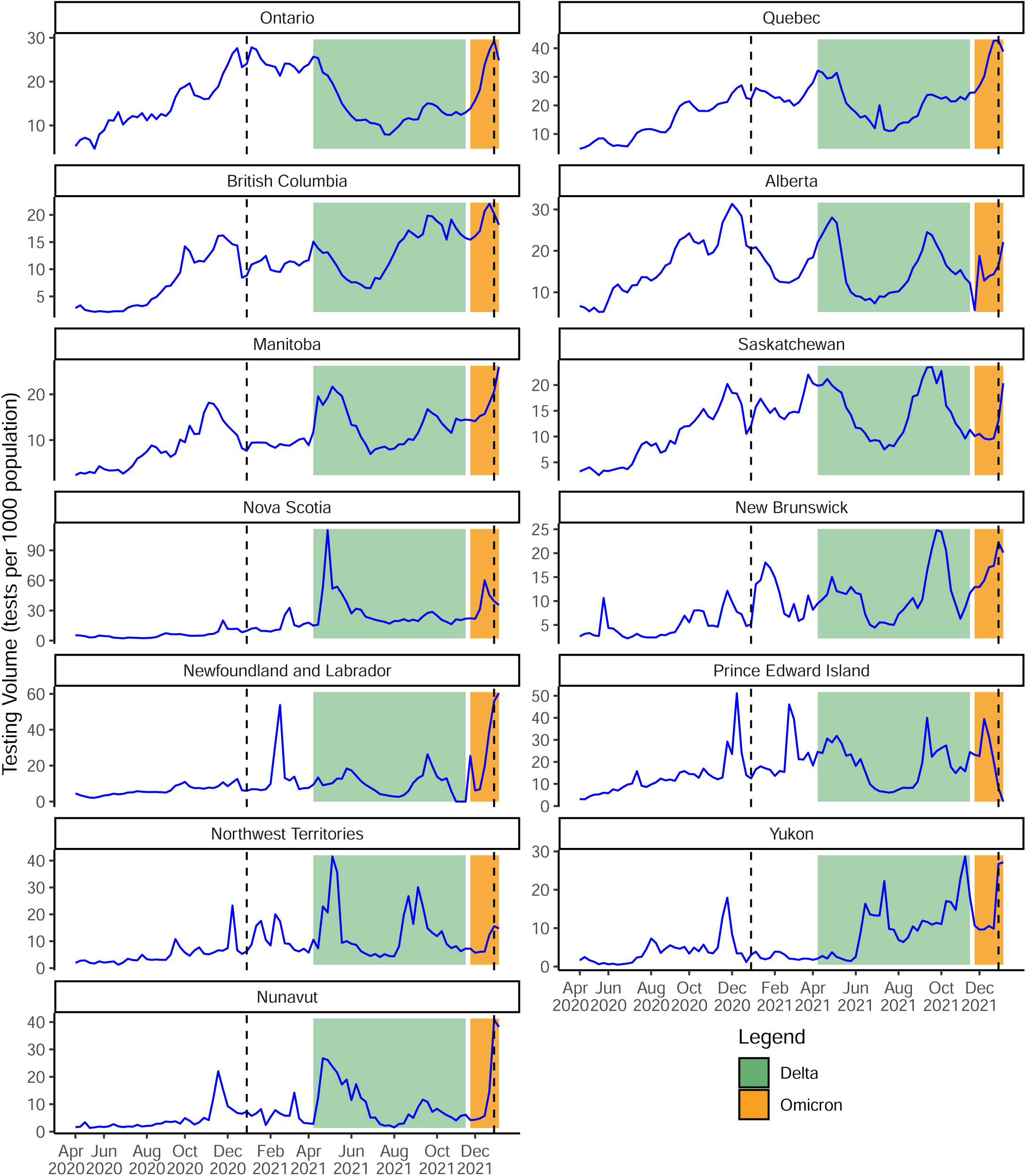
Testing volumes per 1,000 population for each province/territory from 2 Apr 2020 to 6 Jan 2022. The time periods for the two variants of concern Delta and Omicron are depicted with coloured bands, indicating the first observations in Canada. The two vertical dashed lines indicate 1 Jan 2021 and 1 Jan 2022.

**Figure S11:**
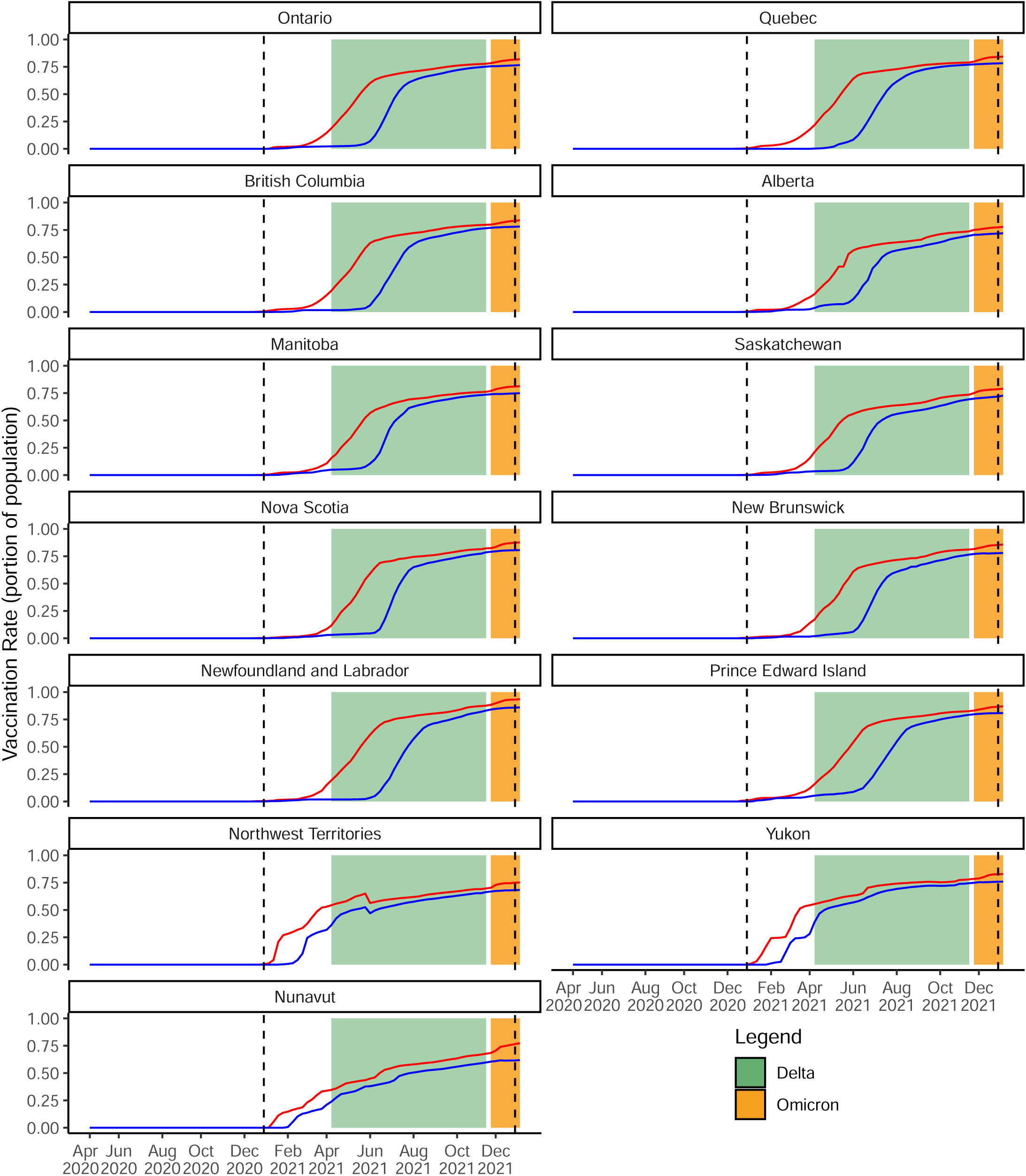
Vaccination rate for each province/territory from 2 Apr 2020 to 6 Jan 2022. The red line indicates at least 1 vaccination dose. The blue line indicates 2 doses. A vaccination rate of 0.5 indicates 50% of the population is vaccinated. The time periods for the two variants of concern Delta and Omicron are depicted with coloured bands, indicating the first observations in Canada. The two vertical dashed lines indicate 1 Jan 2021 and 1 Jan 2022.

**Figure S12:**
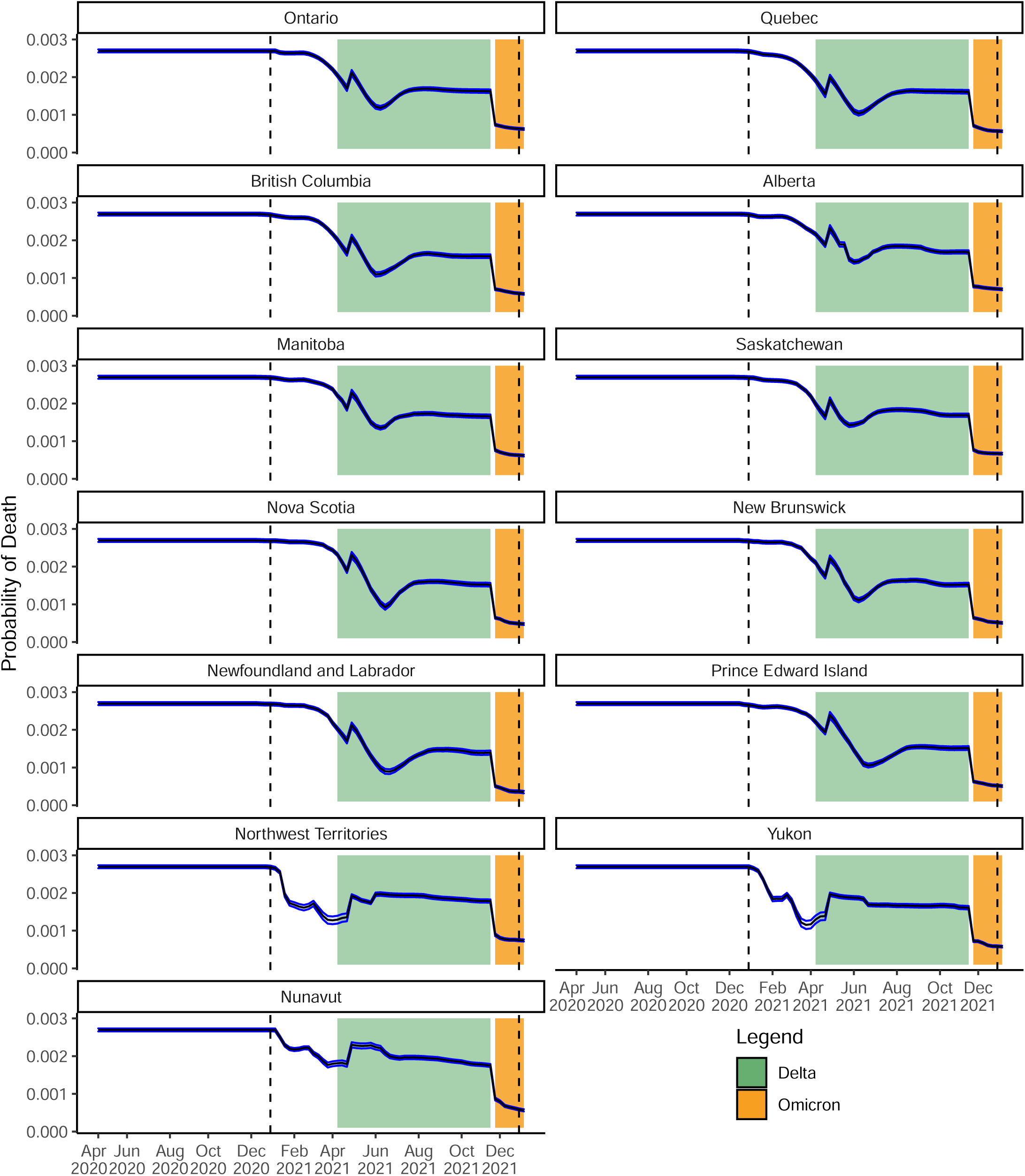
Estimated weekly probability of death (blue) for active COVID-19 cases for each province/territory from 2 Apr 2020 to 6 Jan 2022. The time periods for the two variants of concern Delta and Omicron are depicted with coloured bands, indicating the first observations in Canada. The two vertical dashed lines indicate 1 Jan 2021 and 1 Jan 2022. Bands show 95% credible intervals.

**Figure S13:**
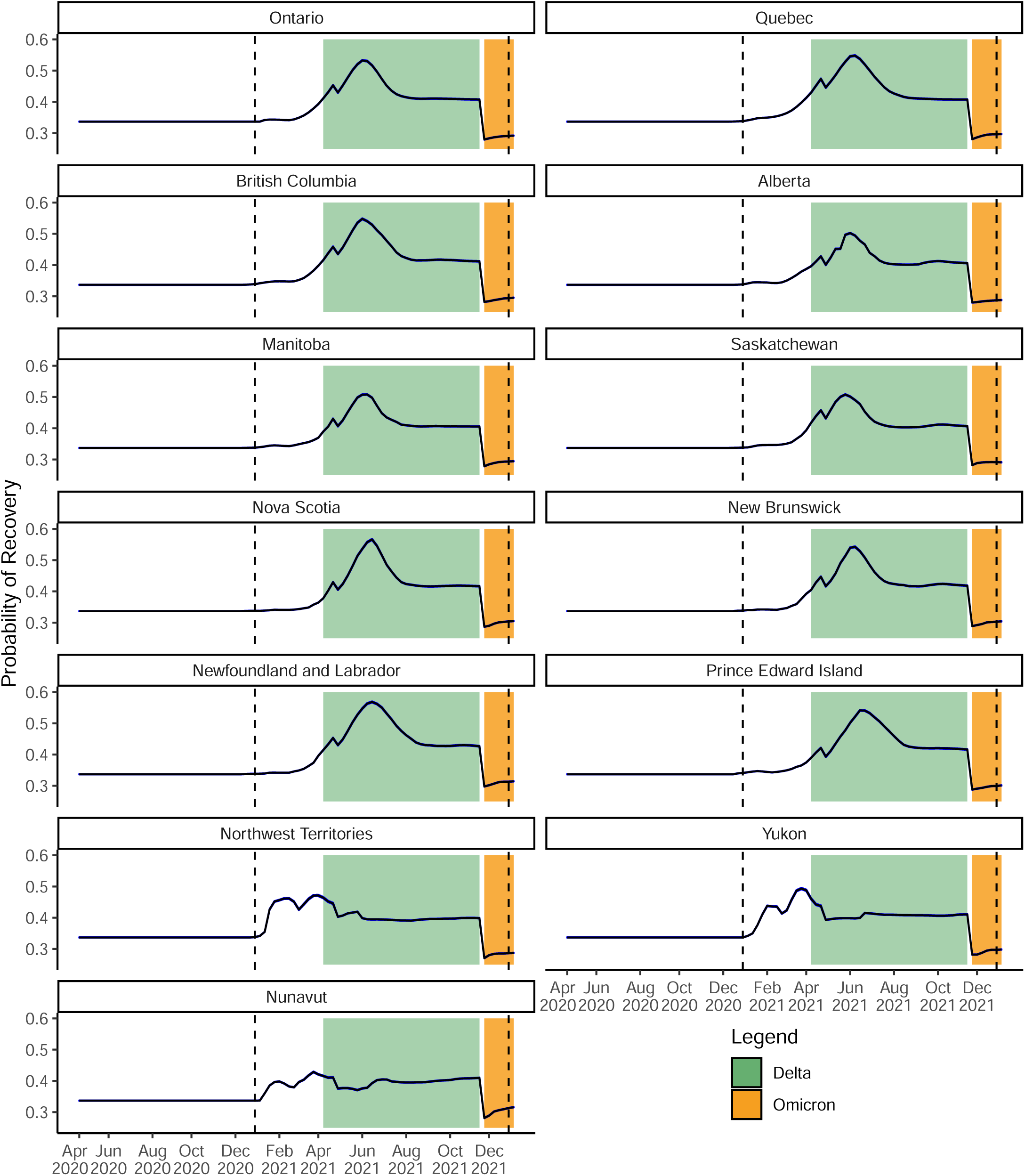
Estimated weekly probability of recovery (blue) for active COVID-19 cases for each province/territory from 2 Apr 2020 to 6 Jan 2022. The time periods for the two variants of concern Delta and Omicron are depicted with coloured bands, indicating the first observations in Canada. The two vertical dashed lines indicate 1 Jan 2021 and 1 Jan 2022. Bands show 95% credible intervals.

**Figure S14:**
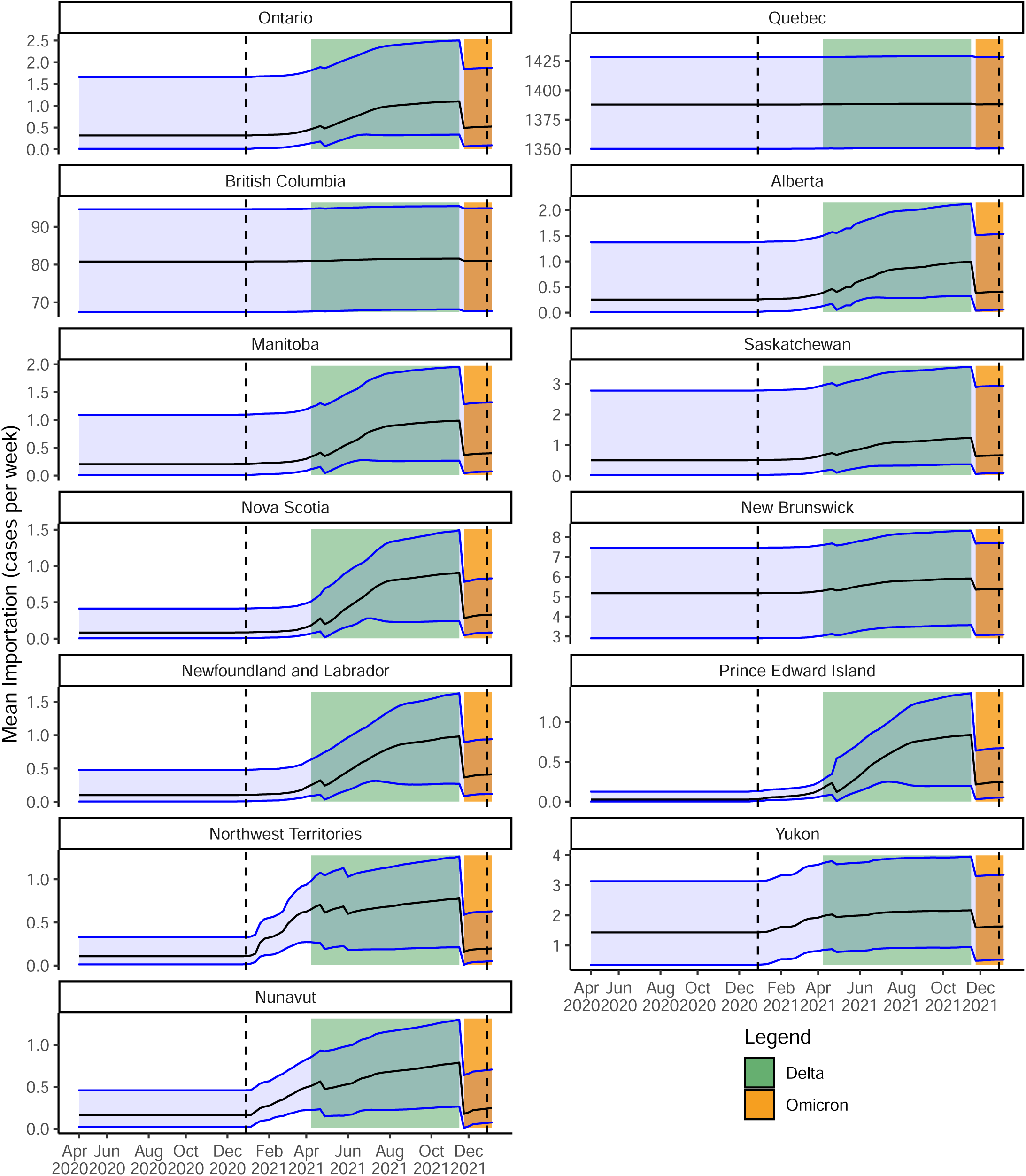
Estimated weekly importation of active COVID-19 cases for each province/territory from 2 Apr 2020 to 6 Jan 2022. The time periods for the two variants of concern Delta and Omicron are depicted with coloured bands, indicating the first observations in Canada. The two vertical dashed lines indicate 1 Jan 2021 and 1 Jan 2022. Bands show 95% credible intervals.

**Figure S15:**
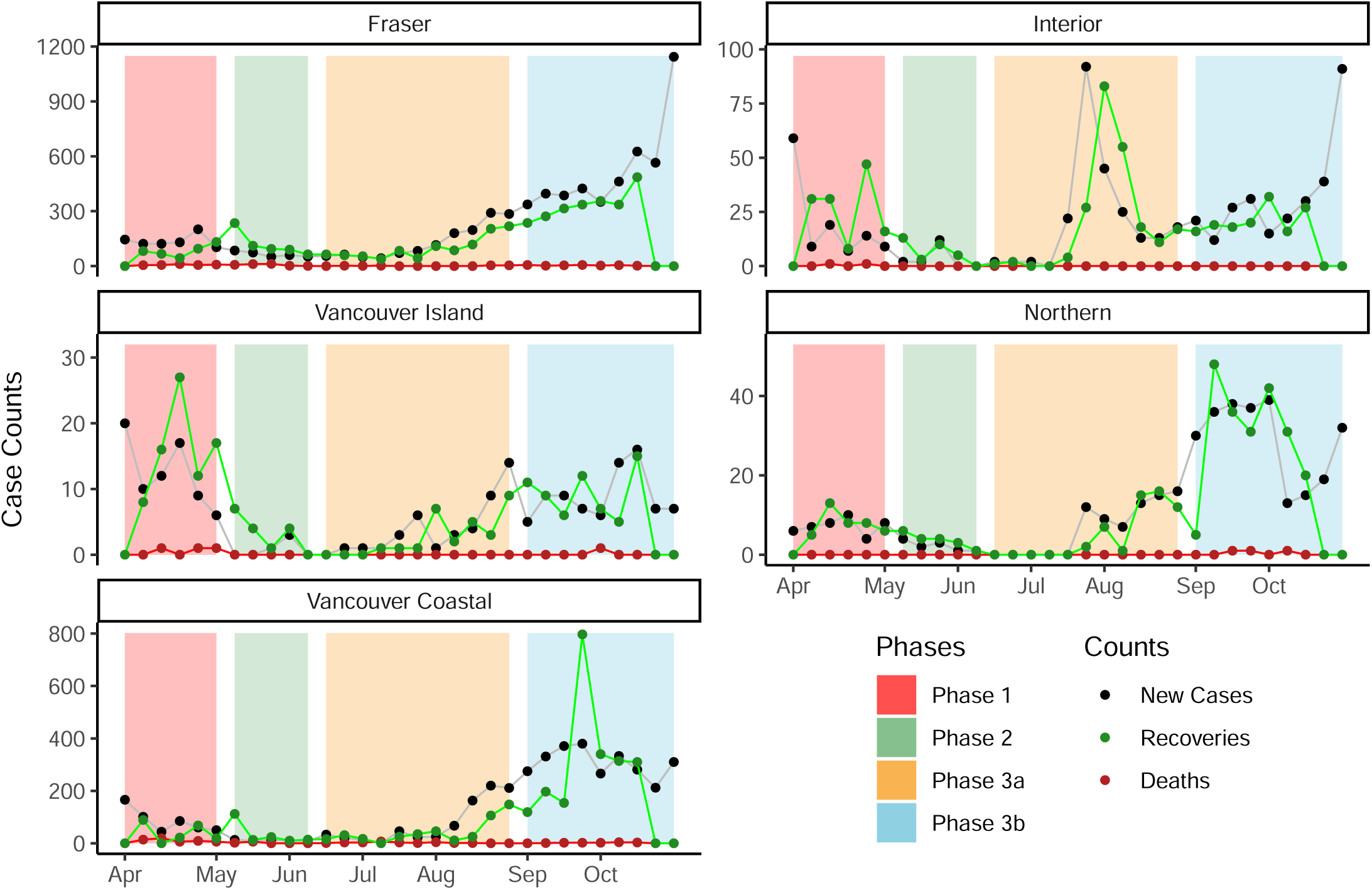
Plots of observed active cases (black), recoveries (green) and deaths (red) in B.C., split by Health Authority Region. Data starts on 2 Apr 2020 (Week 1) and ends on 30 Oct 2020 (Week 31).

**Figure S16:**
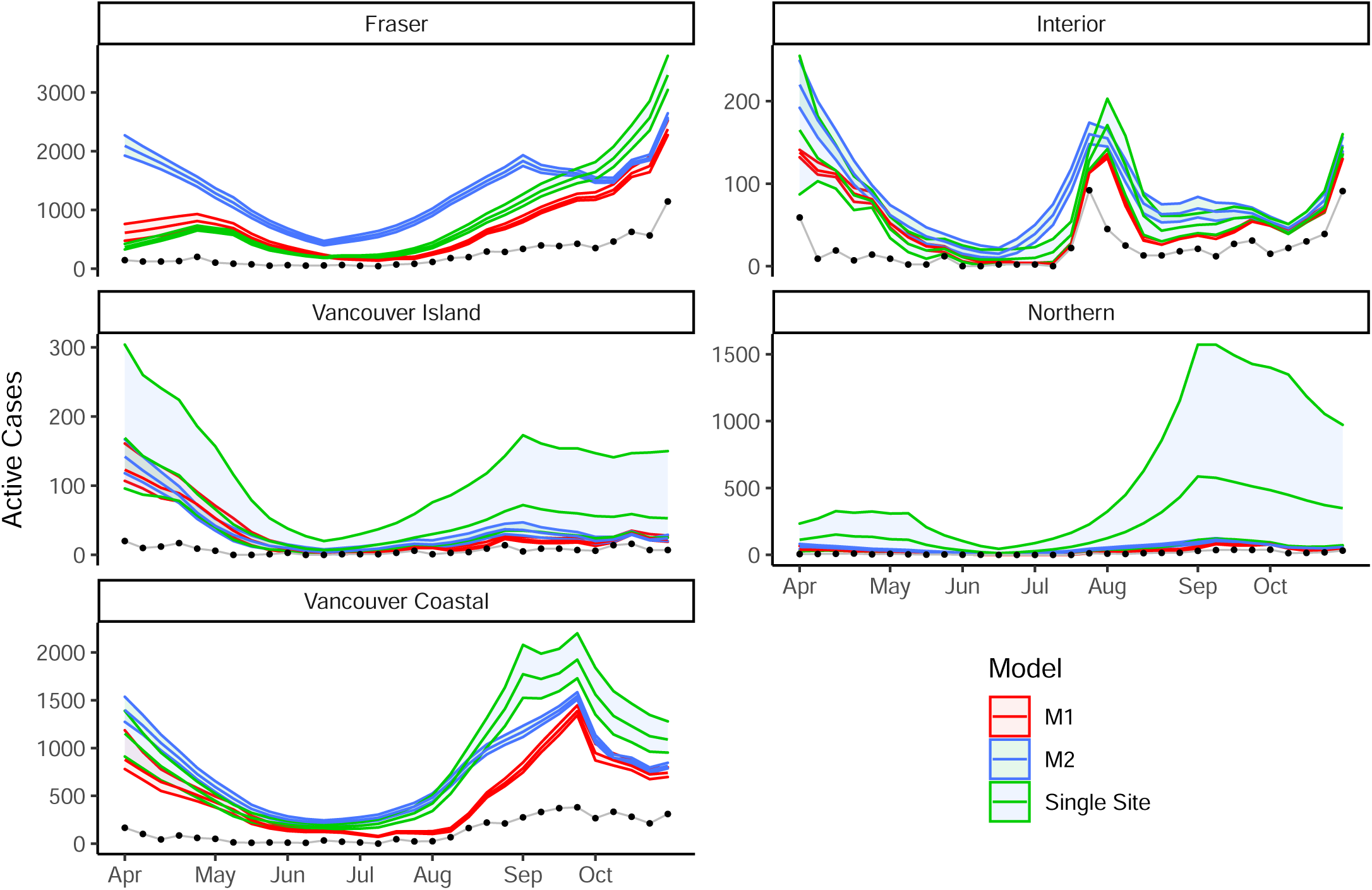
Plots of active cases split by Health Authority Region of B.C. Data starts on 2 Apr 2020 (Week 1) and ends on 30 Oct 2020 (Week 31). The red bands show the 95% credible intervals for total active cases as estimated using model M1, the best fitted model according to WAIC. The blue bands show the 95% credible intervals for total active cases as estimated using model M2, the second best fitted model according to WAIC. The green bands show the 95% credible intervals for total active cases as estimated using the single site models with testing volume as a covariate for detection probability. The black dotted line shows the newly observed active cases each week (according to data).

**Figure S17:**
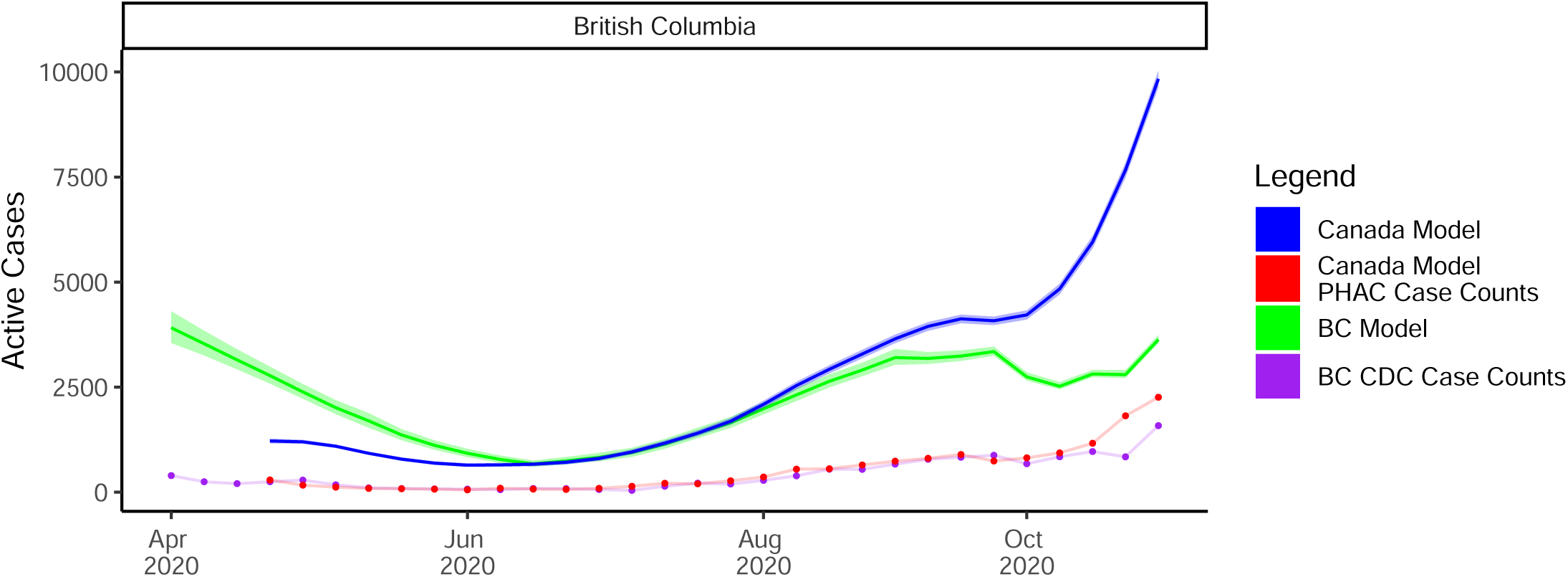
Plots of total active cases for B.C. Data starts on 2 Apr 2020 (Week 1) and ends on 30 Oct 2020 (Week 31). Note that the Canada model data starts on date 23 Apr 2020 (Week 4 of the B.C. only model). The blue bands show the 95% credible intervals for total active cases. Results from the B.C. Health Authority case study, summed over regions, are shown in green, with data shown in purple. Results from the Canada case study, subset to B.C. and Weeks 1 to 31, are shown in blue, with data shown in red. The black dotted line shows the newly observed active cases each week.

### S.6 Supplementary Code

Data and code for the B.C. and Canada-wide case studies are available at https://github.com/mrparker909/COVID-MultiSiteModel2022.git.

